# The International Sexual Health And Reproductive Health Survey (I-SHARE-1): A Multi-Country Analysis of Adults from 30 Countries Prior to and During the Initial COVID-19 Wave

**DOI:** 10.1101/2021.09.18.21263630

**Authors:** Jennifer Toller Erausquin, Rayner K. J. Tan, Maximiliane Uhlich, Joel M. Francis, Navin Kumar, Linda Campbell, Wei-Hong Zhang, Takhona G. Hlatshwako, Priya Kosana, Sonam Shah, Erica M. Brenner, Lore Remmerie, Aamirah Mussa, Katerina Klapilova, Kristen Mark, Gabriela Perotta, Amanda Gabster, Edwin Wouters, Sharyn Burns, Jacqueline Hendriks, Devon J. Hensel, Simukai Shamu, Jenna Marie Strizzi, Tammary Esho, Chelsea Morroni, Stefano Eleuteri, Norhafiza Sahril, Wah Yun Low, Leona Plasilova, Gunta Lazdane, Michael Marks, Adesola Olumide, Amr Abdelhamed, Alejandra López Gómez, Kristien Michielsen, Caroline Moreau, Joseph D. Tucker, I-SHARE research consortium

**Affiliations:** Department of Public Health Education, University of North Carolina Greensboro, Greensboro, USA; Dermatology Hospital of Southern Medical University, Guangzhou, China; University of North Carolina Project-China, Guangzhou, China; Saw Swee Hock School of Public Health, National University of Singapore, Singapore; Department of Psychology, University of Fribourg, Fribourg, Switzerland; Department of Family Medicine, School of Clinical Medicine, University of Witwatersrand, Johanessburg, South Africa; Department of Sociology, Yale University, New Haven, CT, USA; Center for Population, Family, and Health, University of Antwerp, Antwerp, Belgium; Department of Public Health and Primary Care, Faculty of Medicine and Health Sciences, University of Ghent, Ghent, Belgium; School of Public Health, Université Libre de Bruxelles, Brussels, Belgium; Institute of Global Health and Infectious Diseases, University of North Carolina at Chapel Hill, Chapel Hill, NC, USA; Botswana Sexual and Reproductive Health Initiative, Botswana Harvard AIDS Institute Partnership, Gaborone, Botswana; Faculty of Humanities, Charles University, Prague, Czech Republic; National Institute of Mental Health, Klecany, Czech Republic; Department of Family Medicine and Community Health, University of Minnesota Medical School, Minneapolis, MN, USA; Faculty of Psychology, University of Buenos Aires, Buenos Aires, Argentina; Gorgas Memorial Institute for Health Studies, Panama City, Panama; Clinical Research Department, Faculty of Infectious and Tropical Diseases, London School of Hygiene and Tropical Medicine, London, UK; Collaboration for Evidence, Research and Impact in Public Health, School of Population Health, Curtin University, Perth, Australia; Department of Pediatrics, Indiana University School of Medicine, Indianapolis, IN, USA; Department of Sociology, Indiana University Purdue University Indianapolis, Indianapolis, IN, USA; Health Systems Strengthening, Foundation for Professional Development, Pretoria, South Africa; School of Public Health, University of Witwatersrand, Johannesburg, South Africa; Department of Public Health, University of Copenhagen, Copenhagen, Denmark; End FGM/C Centre of Excellence, Amref Health Africa, Nairobi, Kenya; MRC Centre for Reproductive Health, University of Edinburgh, Edinburgh, UK; Department of Psychology, Sapienzo University, Rome, Italy; Ministry of Health Malaysia, Putrajaya, Malaysia; Asia-Europe Institute, Universiti Malaya, Kuala Lumpur, Malaysia; Institute of Public Health, Riga Stradins University, Riga, Latvia; College of Medicine, University of Ibadan, Ibadan, Nigeria; Department of Dermatology, Venereology & Andrology, Sohag University, Sohag, Egypt; Department of Psychology, University of the Republic, Montevideo, Uruguay; Department of Population, Family, and Reproductive Health, Johns Hopkins Bloomberg School of Public Health, Baltimore, MD, USA; Primary Care and Prevention, CESP, INSERM 1018, Villejuif, France

## Abstract

**Background:** The COVID-19 pandemic forced billions of people to shelter in place, altering social and sexual relationships worldwide. In many settings, COVID-19 threatened already precarious health services. However, there is limited evidence to date about changes to sexual and reproductive health (SRH) during the initial wave of COVID-19 disease. To address this gap, our team organized a multi-country, cross-sectional online survey as part of a global consortium.

**Methods:** Consortium research teams conducted online surveys in 30 countries. Sampling methods included convenience, online panels, and population-representative. Primary outcomes included sexual behaviors, partner violence, and SRH service utilization, and we compared three months prior to and three months after policy measures to mitigate COVID-19. We used established indicators and analyses pre-specified in our protocol. We conducted meta-analyses for primary outcomes and graded the certainty of the evidence using Cochrane methods. Descriptive analyses included 22,724 individuals in 25 countries. Five additional countries with sample sizes <200 were included in descriptive meta-analyses.

**Results:** Respondents were mean age 34 years; most identified as women (15160; 66.7%), cis-gender (19432; 86.6%) and heterosexual (16592; 77.9%). Among 4546 respondents with casual partners, condom use stayed the same for 3374 (74.4%) people and 640 (14.1%) people reported a decline. Fewer respondents reported physical or sexual partner violence during COVID-19 measures (1063/15144, 7.0%) compared to the period before COVID-19 measures (1469/15887, 9.3%). COVID-19 measures impeded access to condoms (933/10790, 8.7%), contraceptives (610/8175, 7.5%), and HIV/STI testing (750/1965, 30.7%). Pooled estimates from meta-analysis indicate during COVID-19 measures, 32.3% (95% CI 23.9-42.1) of people needing HIV/STI testing had hindered access, 4.4% (95% CI 3.4-5.4) experienced partner violence, and 5.8% (95% CI 5.4-8.2) decreased casual partner condom use (moderate certainty of evidence for each outcome). Meta-analysis findings were robust in sensitivity analyses that examined country income level, sample size, and sampling strategy.

**Conclusion:** Open science methods are feasible to organize research studies as part of emergency responses. The initial COVID-19 wave impacted SRH behaviors and access to services across diverse global settings.

## Introduction

The COVID-19 pandemic has profoundly disrupted social relationships and health services that are fundamental to sexual and reproductive health.^1^ The initial wave of SARS-CoV-2 infections (COVID-19 disease) forced billions of people worldwide to shelter in place, transforming social and sexual relationships. Entrenched gender inequalities that existed prior to COVID-19 may have been exacerbated during the emergency response,^2^ placing people at increased risk for intimate partner violence (IPV). At the same time, a wide range of essential sexual and reproductive health services were stopped or re-oriented because of the pandemic.^3^ These trends suggest an important question: How have COVID-19 measures impacted sexual and reproductive health outcomes in different settings? Here we define COVID-19 mitigation measures as responses (e.g., non-pharmacological interventions) to slow or halt the spread of the virus within a population, including shelter in place, test and trace, quarantine, and travel restrictions.^4^

Although cities, nations, regions, and the entire world have moved together in altering social lives during the COVID-19 pandemic, there has been substantial variation in COVID-19 disease incidence and responses at the national level. Some countries have imposed less stringent lockdown measures, allowing greater movement between and within cities, while others have instituted more unyielding measures.^5^ Several countries already had infrastructure in place for decentralized sexual and reproductive health services (e.g., HIV self-testing, telemedicine abortion) which compensated for pandemic-related closures of facility-based services during COVID-19.^6^ However, in most countries, COVID-19 further undermined already fragile health infrastructure and health service provision.^7^

Despite the importance of sexual and reproductive health during the initial wave of the COVID-19 pandemic, research in this area is limited.^8,9^ Modeling and other research studies have noted the lack of detailed information about sexual and reproductive health during this period.^10,11^ The lack of standardized survey instruments makes cross-country comparisons more difficult. Most of the sexual and reproductive health research on initial COVID-19 waves has focused on high-income countries,^8^ rather than examining broader regional and global trends. Few studies to date have included low and middle-income countries.^9^ At the same time, the global pandemic has accelerated open science and spurred new forms of collaboration.

Our team organized a cross-sectional multi-country study called “International Sexual Health And REproductive Health during COVID-19” (I-SHARE-1).^12^ The I-SHARE project convened a group of sexual and reproductive health researchers to administer a common survey instrument in respective countries as an online survey.^13^ Any research team could join and teams were identified through an earlier UNDP/UNFPA/UNICEF/WHO/World Bank Special Programme of Research, Development and Research Training in Human Reproduction (HRP) crowdsourcing open call^12^ and a related open call through affiliates of the Academic Network for Sexual and Reproductive Health and Rights (ANSER). The purpose of this multi-country study was to better understand sexual and reproductive health prior to and during the first wave of the COVID-19 pandemic in respective countries.

## Methods

A more detailed description of survey methods can be found in the protocol.^12^ The primary aims of the study were to examine changes in sexual behaviors (sex frequency and condomless sex), intimate partner violence, and utilization of sexual and reproductive health services during COVID-19 measures using a cross-sectional survey. Secondary study aims were to examine changes in HIV/STI testing, harmful cultural practices (e.g., female genital mutilation/cutting and child marriage), mental health, and food security. Each country adjusted the questionnaire based on country-level priorities, opportunities, and needs. The consortium recommended a sample size of at least 200, but precise sample size calculations were made by each country’s research team. We used an open science approach in organizing this study. This approach included allowing any interested research team to join the project, facilitating collaboration between sites, leveraging open-access software, and prioritizing open access outputs.

### Recruitment and Participants

Participants were recruited through an online survey link that was distributed through local, regional, and national networks. Recruitment used social media (26 studies), partner organizations (20 studies), paid social media advertising (11 studies), university websites (10 studies), telephone interviews (4 studies), television or newspapers (3 studies). Thirty countries implemented the study, including Argentina, Australia, Botswana, Canada, China, Colombia, Czech Republic, Denmark, Egypt, France, Germany, Italy, Kenya, Latvia, Lebanon, Luxembourg, Malaysia, Mexico, Moldova, Mozambique, Nigeria, Panama, Portugal, Singapore, South Africa, Sweden, Spain, Uganda, United States, and Uruguay (Supplemental Table 1). A total of twenty-three studies used convenience sampling (Australia, Canada, Colombia, China, Czech Republic, Egypt, France, Germany, Italy, Latvia, Panama, Portugal, Luxembourg,

Mexico, Malaysia, Moldova, Mozambique, Nigeria, Singapore, South Africa, Spain, Uruguay, USA), six studies used online panels (Sweden, Botswana, Uganda, Lebanon, Kenya, Argentina), and two used population-based methods (Czech Republic, Denmark). Consortium members in the Czech Republic conducted two separate studies (one using a convenience sample and one using a population-based sample), and thus a total of 31 studies among 30 countries were reported. Eligible participants were age 18 years or older (or younger if the country’s Institutional Review Board and ethical regulation permitted it and the in-country lead ensured appropriate procedures), resided in the respective participating country, were capable of reading and understanding the survey language, could access an online survey, and were willing to provide informed consent.

### Survey development

The partners collaboratively developed the survey instrument based on existing items from a recent WHO survey instrument intended for global use,^14^ other existing tools, and items adapted for COVID-19. The survey included the following sections: sociodemographic characteristics; compliance with COVID-19 measures; couple and family relationships; sexual behavior; contraceptive use and barriers to access; access to reproductive healthcare; abortion; sexual violence and IPV; HIV/STI testing and treatment; female genital mutilation/cutting and early/forced marriage (optional); mental health (optional); and food insecurity (optional) (Supplemental Tables 2 and 3). The time periods for pre-COVID-19 and during initial COVID-19 measures were specified for each of the studies based on the in-country team.

The lead organization in each country selected networks to disseminate the survey link, and it was primarily distributed through email lists, local partner organizations affiliated with ANSER, other sexual and reproductive health networks, and social media links. The survey took most participants 20-30 minutes to complete.

Each country had a research team that led the country’s ethical review, translation and survey administration while providing support and organization for the multinational study. The survey was available in the official language of the country and other relevant languages. In total, the survey was translated into 21 languages. In most participating countries, CAPTCHA or other fraud protection methods were included to prevent more than one response from a single IP address.

The survey included several potentially sensitive questions including items about sexuality, gender identity, sexual behavior, abortion, and IPV. Participants could stop the survey at any point or leave out questions they did not want to answer. Participating country institutions signed data sharing agreements for cross-country analysis. Resources on country-specific referral pathways for IPV, sexual health services, and reproductive health services were provided at the end of the survey. All data were de-identified before multi-country analyses.

### Data analysis

Multi-country analysis was undertaken for countries that met specific pre-specified criteria. Each country was required to have obtained Institutional Review Board approval from a local ethics authority, locally translated and field-tested the instrument, described the sampling methodology, and obtained responses from at least 200 participants. A minimum threshold of 200 participants was used because small samples may be more likely to be biased and have higher heterogeneity. We examined the effect of including all data empirically using a sensitivity analysis. We did not weight our estimates because most countries did not use a probability sample. We conducted descriptive meta-analysis to assess the effect of study characteristics and setting and more accurately estimate the prevalence of our key outcomes across multiple countries.

First, we ran descriptive statistics on using the main data set of 25 countries to assess patterns in respondent sociodemographic characteristics and to assess the primary outcomes prior to and during COVID-19 measures. We used the Oxford indices to assess the stringency of COVID-19 measures in each country, based on the mean value across the days when the survey was open. We used the Appraisal Tool for Cross-Sectional Studies (AXIS) to assess risk of bias.^15^ Second, we conducted a meta-analysis for all 30 countries on the prevalence of reported hindered access to HIV/STI testing, IPV during COVID-19 measures, and decreased condom use with casual partners. We used meta-analysis because this provided a mechanism to assess risk of bias of individual studies and consider the strength of the evidence. Tests for heterogeneity were applied using I^2^ statistics.^16^ We also adopted the GRADE (Grading of Recommendations, Assessment, Development and Evaluations) framework to rate the quality of evidence presented in our meta-analysis.^17^ Furthermore, we conducted sensitivity analyses that separated primary outcomes based on country income level (low and middle-income countries compared to high-income countries), sample size (less than 200 or more), and sampling strategy (convenience compared to online panel or population-representative). All analyses were carried out using Stata version 14, and missing data were treated by pairwise deletion (available-case analysis).

## Results

### Results of descriptive analysis

Twenty-five of the 30 countries that joined the I-SHARE study (Figure 1) met all study criteria, including recruiting a minimum of 200 participants. Five countries (Mozambique, Canada, Egypt, Lebanon, and South Africa) had fewer than 200 participants and were excluded from descriptive analyses. The majority of countries across all four geographic regions implemented all survey components, except FGM and early marriage (Supplemental Table 2). Abortion and mental health components were excluded in 2 and 3 countries, respectively.

**Figure 1.**
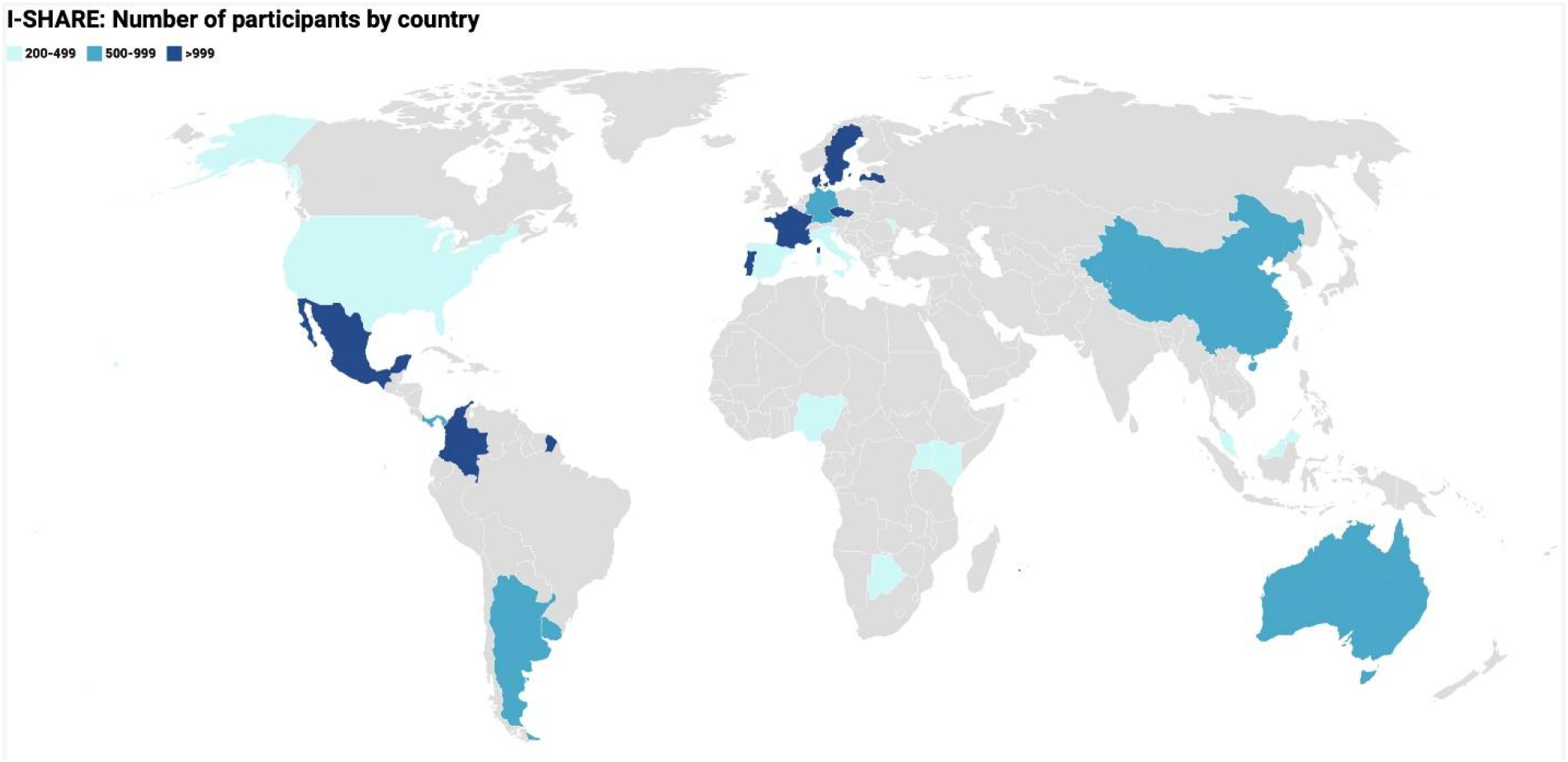
World map with 25 countries included in ISHARE-1 highlighted.

Among the 25 included countries, 14 were high-income countries, eight were upper-middle-income, two were lower-middle-income, and one was low-income (see Supplemental Table 1). There was a wide geographic distribution, with eleven countries in Europe, six in the Americas, four in Asia and Oceania, and four in Africa. In terms of severity of COVID-19 measures, twelve countries were moderate or “middle stringency” on the Oxford Stringency Index and 13 were high stringency. There was variation in the total sample size recruited from each country, with eight countries having more than 1000 participants, eight having 500-999 participants and nine having between 200 and 499 participants.

As shown in Table 1, two-thirds (66.7%) of participants were women, and over 8 in 10 participants (86.6%) were cis-gender. About 78% of participants were heterosexual. Most participants (44.6%) were 18-29 years old, followed by those 30-39 (26.9%) and 40-49 (14.4%) years old. Few participants (2.9%) were 70 years or older. More than half (55.9%) of participants reported having completed a college degree. There was diversity in reported socioeconomic position of the household relative to others in their country, with most participants (38.4%) indicating that their household was in the 5^th^ or 6^th^ highest income group out of 10 in their country. Nearly three-quarters (74.0%) of participants reported living in an urban or semi-urban area.

**Table 1.** Sociodemographic characteristics of participants in the I-SHARE multi-country survey, 2020-2021 (n = 22,724).

| *Variable* | *Level* | **n** | **%** |
| --- | --- | --- | --- |
| *Sex assigned at birth* | Female | 15160 | 66.7 |
|  | Male | 7505 | 33.0 |
|  | Another sex* | 52 | 0.2 |
|  | Total | 22717 | 100 |
| Gender | Cisgender | 19432 | 86.6 |
|  | Non-cisgender | 2672 | 11.9 |
|  | Another gender | 338 | 1.5 |
|  | Total | 22442 | 100 |
| *Sexual Orientation* | Heterosexual | 16592 | 77.9 |
|  | Bisexual | 1823 | 8.6 |
|  | Gay | 818 | 3.8 |
|  | Asexual | 629 | 3.0 |
|  | Questioning or unsure | 446 | 2.1 |
|  | Other | 351 | 1.7 |
|  | Lesbian | 315 | 1.5 |
|  | Pansexual | 315 | 1.5 |
|  | Total | 21289 | 100 |
| *Age group in years* | 18-29 | 10135 | 44.6 |
|  | 30-39 | 6109 | 26.9 |
|  | 40-49 | 3268 | 14.4 |
|  | 50-59 | 1644 | 7.2 |
|  | 60-69 | 916 | 4.0 |
|  | 70+ | 652 | 2.9 |
|  | Total | 22724 | 100 |
| *Education* | No formal education | 102 | 0.5 |
|  | Some or completed primary school | 944 | 4.2 |
|  | Some or completed secondary school | 4717 | 20.8 |
|  | Some college or university | 3457 | 15.3 |
|  | Completed college or university | 12619 | 55.7 |
|  | Other | 803 | 3.6 |
|  | Total | 22,642 | 100 |
| *Relative household socioeconomic position (1-10)\*\** | Lower position (1-2) | 2227 | 11.1 |
|  | 3-4 | 4319 | 21.5 |
|  | 5-6 | 7712 | 38.4 |
|  | 7-8 | 4327 | 21.6 |
|  | Higher position (9-10) | 1486 | 7.4 |
|  | Total | 20071 | 100 |
| *Urban/Rural* | Urban or semi-urban | 15722 | 74.0 |
|  | Rural or semi-rural | 4710 | 22.2 |
|  | Other | 809 | 3.8 |
|  | Total | 21241 | 100 |
| *Relationship Status* | Single, never had partner | 2113 | 9.3 |
|  | Single, ever had partner | 4268 | 18.8 |
|  | In a relationship, not cohabiting | 4354 | 19.2 |
|  | Not married, cohabiting | 4349 | 19.1 |
|  | Legally married, cohabiting | 5753 | 25.3 |
|  | Legally married, not cohabiting | 1083 | 4.8 |
|  | Separated or divorced | 894 | 3.9 |
|  | Widowed | 178 | 0.8 |
|  | Other | 285 | 1.3 |
|  | Total | 22724 | 100 |
| *Current pregnancy situation* | Currently pregnant | 514 | 3.7 |
|  | Currently trying to become pregnant | 835 | 6.1 |
|  | Recently had a baby | 432 | 3.1 |
|  | Not trying | 10377 | 75.2 |
|  | Cannot have children | 1584 | 11.5 |
|  | Other | 60 | 0.4 |
|  | Total | 13802 | 100 |
| *Sexual Activity Frequency (Steady Partner)* | Never | 811 | 5.3 |
|  | Monthly or less | 2366 | 15.4 |
|  | 2-4 times a month | 5758 | 37.6 |
|  | 2-3 times a week | 4583 | 29.9 |
|  | 4 or more times a week | 1802 | 11.8 |
|  | Total | 15320 | 100 |
| *Sexual Activity Frequency (Casual Partner)* | Never | 15655 | 75.9 |
|  | Monthly or less | 3181 | 15.4 |
|  | 2-4 times a month | 1375 | 6.7 |
|  | 2-3 times a week | 316 | 1.5 |
|  | 4 or more times a week | 96 | 0.5 |
|  | Total | 20623 | 100 |
| *Sex Life Satisfaction (Before COVID-19)* | Very satisfied | 7535 | 36.6 |
|  | Somewhat satisfied | 8026 | 39.0 |
|  | Neutral | 216 | 1.1 |
|  | Not very satisfied | 3431 | 16.7 |
|  | Not at all satisfied | 1382 | 6.7 |
|  | Total | 20590 | 100 |
| *Sex Life Satisfaction (During COVID-19)* | Very satisfied | 5484 | 26.7 |
|  | Somewhat satisfied | 6738 | 32.8 |
|  | Neutral | 202 | 1.0 |
|  | Not very satisfied | 4788 | 23.3 |
|  | Not at all satisfied | 3353 | 16.3 |
|  | Total | 20565 | 100 |

\*This included individuals whose sex at birth was not a male or female.\*\* Oxford Stringency Index: a measure to record the strictness of lockdown policies based on indicators such as school and workplace closure, and travel bans, rescaled to a value from 0 to 100 (100 = strictest).
Note: Household socioeconomic status and relationship status were not mutually exclusive and participants could choose more than one.

The lower panel of Table 1 presents relationship status and sexual frequency, and sexual satisfaction in the three months before and during COVID-19 measures. There were a variety of relationship types reported, with 43.4% in a cohabiting relationship. Among sexually experienced participants, most (75.2%) were not pregnant and not trying to become pregnant. Among those with a steady partner, 37.6% reported having sex with that partner 2-4 times a month, and another 29.9% reported 2-3 times a week. Among those with a casual partner, the most commonly reported frequency of sex with that partner was monthly or less (15.4%). Most participants (75.6%) reported being somewhat satisfied or very satisfied with their sex life before COVID-19, but this proportion had fallen (to 59.4%) during COVID-19 in the same participants.

In terms of compliance with COVID-19 measures (Supplemental Table 5), 58.9% of participants reported they had followed measures a lot. The majority (76.6%) had never been in isolation due to their own symptoms or close contact with someone with COVID-19, and two-thirds (66.2%) had never been tested for COVID-19. Although 62.2% of participants said that their household socioeconomic status stayed the same during the COVID-19 pandemic, about one-third (32.0%) reported their household economic situation worsened.

Table 2 shows our key study outcomes before and during COVID-19. Condom use “always” or “most of the time” with steady partners (62.5%) and with casual partners (63.8%) was relatively high prior to COVID-19 measures. Although most participants perceived their condom use stayed the same during COVID-19 measures (74.4% with casual partners and 86.9% with steady partners), 14.1% of participants with casual partners (and 10.4% of those with steady partners) reported their condom use with those types of partners decreased during COVID-19 measures. Regarding physical or sexual violence, 9.3% reported experiencing one or more types of violence prior to COVID-19, and a slightly lower proportion (7.0%) reported experiencing these types of violence during COVID-19 measures. Additional analyses showed that among those reporting no prior physical or sexual violence from a partner, 1.4% reported experiencing violence during COVID-19 measures; among those who did report prior physical or sexual violence from a partner, 67.9% reported experiencing violence during COVID-19 measures.

**Table 2.** Key outcomes 3 months before and during COVID-19 social distancing measures in the 25 I-SHARE countries with ≥200 respondents, 2020

|  |  | **N** | **%** | **95% CI** |
| --- | --- | --- | --- | --- |
| **Condom Use with Steady Partners (Before)** |  | N=3,282 |  |  |
|  | Always or Most of the Time | 2,050 | 62.46 | (60.78-64.12) |
|  | Sometimes/Rarely/Never | 1,232 | 37.54 | (35.88-39.22) |
| **Condom Use with Casual Partners (Before)** |  | N=4,383 |  |  |
|  | Always or Most of the Time | 2,795 | 63.77 | (62.33-65.19) |
|  | Sometimes/Rarely/Never | 1,588 | 36.23 | (34.81-37.67) |
| **Perceived Changes to Condom Use with Steady Partners (During)** |  | N=12,183 |  |  |
|  | Decreased | 1,262 | 10.36 | (9.82-10.91) |
|  | Stayed the same | 10,588 | 86.91 | (86.29-87.50) |
|  | Increased | 333 | 2.73 | (2.45-3.04) |
| **Perceived Changes to Condom Use with Casual Partners (During)** |  | N=4,546 |  |  |
|  | Decreased | 640 | 14.08 | (13.08-15.12) |
|  | Stayed the same | 3,374 | 74.22 | (72.92-75.49) |
|  | Increased | 532 | 11.70 | (10.78-12.67) |
| **Any Physical or Sexual Violence from Partner (Before)** |  | N=15,887 |  |  |
|  | No | 14,418 | 90.75 | (90.29-91.20) |
|  | Yes | 1,469 | 9.25 | (8.80-9.71) |
| **Any Physical or Sexual Violence from Partner (During)** |  | N=15,144 |  |  |
|  | No | 14,081 | 92.98 | (92.56-93.38) |
|  | Yes | 1,063 | 7.02 | (6.62-7.44) |
| **Among those reporting no prior physical or sexual violence from a partner, 1.4% reported experiencing violence during COVID-19 measures. Among those who did report prior physical or sexual violence from a partner, 67.9% reported also experiencing violence during COVID-19 measures.** |  |  |  |  |
| **COVID-19 measures made it more difficult to access condoms** |  | N=10,790 |  |  |
|  | No | 9,857 | 91.35 | (90.80-91.87) |
|  | Yes | 933 | 8.65 | (8.12-9.19) |
| **COVID-19 measures stopped or hindered you from seeking contraceptives** |  | N=8,175 |  |  |
|  | No | 7,565 | 92.54 | (91.95-93.10) |
|  | Yes | 610 | 7.46 | (6.90-8.05) |
| **COVID-19 measures stopped or hindered you from seeking or obtaining an abortion\*** |  | N=150 |  |  |
| **\* among those reporting being in need of abortion during COVID-19** |  |  |  |  |
|  | No | 104 | 69.33 | (61.29-76.59) |
|  | Yes | 46 | 30.67 | (23.41-38.71) |
| **COVID-19 measures stopped or hindered you from accessing a test for HIV or STIs\*** |  | N=1,965 |  |  |
| **\* among those reporting wanting an HIV or STI test** |  |  |  |  |
|  | No | 1,215 | 61.83 | (59.64-63.99) |
|  | Yes | 750 | 38.17 | (36.01-40.35) |

For sexual and reproductive health care access, we first examined condom access. About 9% of participants indicated that COVID-19 measures made it more difficult to access condoms. A slightly smaller proportion (7.5%) reported that COVID-19 measures stopped or hindered contraceptive access. Nearly one-third (30.7%) of participants who reported needing abortion services during COVID-19 reported that COVID-19 measures stopped or hindered them from seeking or obtaining this service. In addition, 38.2% of participants that needed HIV/STI testing reported that COVID-19 measures stopped or hindered them from accessing HIV or STI testing.

### Results of meta-analyses

Meta-analyses using data from all 30 countries indicated substantial heterogeneity at the country level for all outcomes, including hindered access to HIV/STI testing (P=.000, I^2^=89.9%), IPV experienced during COVID-19 measures (P=.000, I^2^=95.5%), and condom use during COVID-19 measures (P=.000, I^2^=95.5%). Pooled estimates suggest that 32.3% (95% CI 23.9 – 42.1%) of people needing HIV/STI testing had hindered access to HIV/STI testing (Supplemental Figures 1-3). Approximately 4.4% (95% CI 3.4 - 5.4%) of people experienced physical or sexual violence (Supplemental Figures 4-6) during COVID-19 measure. Finally, 5.8% (95% CI 5.4 – 8.2%) of people reported a decrease in condom use with sexual partners during COVID-19 measures (Supplemental Figures 7-9).

Risk of bias assessment for the studies in I-SHARE indicated that, in general, study procedures of all studies were largely justified, appropriate, and adequately described (Supplemental Table 5). The convenience sampling methods used by most countries introduced bias. In addition, response rates raised concerns about non-response bias and information about non-responders was not available.

The GRADE framework was used to assess the quality of evidence for each of the three meta-analysis outcomes (Supplemental Table 6). Each of the three main findings was associated with a moderate certainty of evidence. Observational studies in general begin at a low quality of evidence; while there were risks of bias due to convenience sampling, we rated the quality of our evidence upwards due to the large effect size for the outcome of hindered access to HIV/STI testing, and the large sample size of the study across all outcomes.

## Discussion

Our study findings provide important insights into sexual and reproductive health during the initial COVID-19 wave in diverse global settings. Our data suggest that condomless sex with casual partners did not substantially change with the introduction of COVID-19 measures. Experiences of intimate partner violence may have decreased during COVID-19 measures compared to prior to the pandemic. Among the health services we examined, there were marked decreases in access to HIV/STI testing and abortion services.

We found that condomless sex was similar during COVID-19 measures compared to the pre-COVID-19 period for many respondents. Approximately 74-87% of people reported that condom use with a steady and/or casual partner stayed the same during these two periods. Maintenance of pre-COVID-19 condom use behavior is consistent with observational studies from sex workers and ethnic and racial minority groups.^18,19^ Given that COVID-19 introduced many new infectious disease risks, some individuals may have been less likely to engage in risky sexual behaviors.^20^ Only 8.7% of the sample noted problems accessing condoms. The COVID-19 environment did not appear to substantially alter individual decisions about whether to use a condom.

Our results suggest a modest decrease in sexual and physical partner violence during COVID-19 measures compared to the pre-COVID period. Although there was concern about COVID-19 exacerbating intimate partner violence,^2^ data on intimate partner violence during the pandemic have been mixed. Some studies suggest increased intimate partner violence during COVID-19 measures,^21,22^ while others found decreases.^23^ Other research has shown that IPV may increase after a natural disaster,^24^ indicating a need for follow up studies to see if IPV worsened as the COVID-19 pandemic continued beyond the initial wave that we examined in this study.

Our study also indicates that COVID-19 measures interrupted access to HIV/STI testing and abortion services. This finding is consistent with other studies observing interruptions in HIV/STI testing^25,26^ and abortion services.^27^ Decentralized testing approaches using STI self-collection and HIV self-testing^28^ have alleviated some of the gaps in diagnostic service provision during COVID-19. However, despite strong evidence that telemedicine is safe and effective for providing medical abortion services, several countries further restricted abortion services during the initial wave of the COVID-19 pandemic.^29^ More research and advocacy are needed to support abortion services during pandemics and similar circumstances.

Our study has several limitations. First, this was an online survey organized during COVID-19 measures, introducing risk for selection bias. Although there is no guideline for conducting online surveys, we used several strategies to limit bias, including the use of online panels, partnerships with organizations for sample recruitment, review of analytics, and prespecified analysis plans.^13^ Second, although we were able to capture data from different times during the COVID-19 epidemic, this was a series of retrospective cross-sectional studies, and we did not capture how sexual behaviors and access evolved over the course of the pandemic. A follow-up survey in selected countries is now underway. Third, our sample included more women, people with higher education, and people living in high-income countries compared to populations in respective countries. At the same time, data from one of the convenience samples included in this analysis suggested that the convenience sample included similar proportions of adults within subnational geographic areas compared to census data.^30^ Fourth, our study had fewer studies from low-income countries which may have been due to later COVID-19 initial waves and less capacity for research alongside the pandemic. At the same time, our main findings were robust when stratifying based on country income level.

Although COVID-19 measures made it more difficult to obtain population-representative samples, we organized a multi-country analysis of data from 30 countries. Several studies have noted that online surveys may be particularly useful for collecting information about sensitive sexual behaviors compared to in-person survey methods.^13^ Strengths of this study include the inclusive open science approach, the harmonization of key sexual health variables across countries, and the geographic diversity.

The use of meta-analysis methods was a key factor in mitigating risks of bias in our study. Pooled estimates of key outcomes reported in this study generated through meta-analysis provided more conservative estimates of our key study outcomes than our descriptive findings, thus mitigating bias in the varying sampling strategies across countries. Sensitivity analyses revealed differences in proportions based on country income level and sample size for experiencing IPV during the COVID-19 measures, while differences in proportions based on country income level and sampling strategy were observed for decreased condom use during COVID-19 measures. Differences in country-level income and sampling strategies do not have any bearing on the presentation of our descriptive findings but offer insight into country-level variations for these outcomes. However, because we omitted countries with sample sizes of less than 200 in our descriptive sample, and those countries omitted had a generally lower level of IPV experienced during COVID-19 measures compared to other countries in the sample, our pooled estimate for the proportion of individuals experiencing IPV may overestimate this outcome.

This study has implications for research and policy. From a research perspective, this underscores the need for sexual behavior, IPV, and reproductive health service access research in emergency settings. Given the heterogeneity in study outcomes, multi-national studies should consider using methods that account for clustering (e.g., multilevel modeling). From a policy perspective, our data suggest the need for expanded use of decentralized sexual and reproductive health interventions that could be implemented in emergency settings (e.g., self-testing, self-collection, telemedicine abortion). The results from country-level data have already helped to inform COVID-19 related sexual and reproductive health policies in several countries, including Latvia, Czech Republic, Panama, Singapore, Uruguay, and Portugal.

Finally, the open science methods used in this study point towards new frameworks for global health collaboration. We organized a survey in thirty diverse settings during a pandemic, despite not having a central funding source or a COVID-19-specific organizational remit. This suggests the feasibility of grounds-up organized multi-country studies focused on sexual and reproductive health.

## Data Availability

Sharing of data for multi-country comparisons is governed by data sharing agreements. Data can only be shared within the I-SHARE research consortium. In-country leads maintain control of individual country data and decisions about country-level data access are made solely by them.

## Acknowledgement

The authors would like to thank the I-SHARE research consortium.

## Author Contributions

RT, JTE, KM, and JT developed the initial idea. RT and JTE led the data analysis with the data analysis subgroup who included MU, KM, EW, TH, SS, MM, JT, WHZ, AM and JF. TH, PK, SS, LC, EB, LR were part of the digital working group that programmed surveys. KK, KM, AG, SB, DH, SS, JS, TE, CM, SE, WL, LP, GL, AO, and CM were all country leads on surveys and led field testing, translation, ethical review applications, and survey implementation at the country level. KM and JT were coordinators for multi-country analysis. All authors read and approved the final version that was submitted.

## Supplemental Tables

**Supplemental Table 1.** I-SHARE Countries with greater than 200 participants (n = 25) in 2020-2021.

|  |  |  |  |
| --- | --- | --- | --- |
| *Country Income Group*<sup>1</sup> | Low-income countries | 1 | Uganda |
|  | Lower middle-income countries | 2 | Kenya, Nigeria |
|  | Upper middle-income countries | 8 | Argentina, Botswana, China, Colombia, Malaysia, Mexico, Moldova (Republic of), Panama |
|  | High income countries | 14 | Australia, Czech Republic, Denmark, France, Germany, Italy, Latvia, Luxembourg, Portugal, Singapore, Spain, Sweden, United States, Uruguay |
| *Region* | Americas | 6 | Argentina, Colombia, Mexico, Panama, United States, Uruguay |
|  | Africa | 4 | Botswana, Kenya, Nigeria, Uganda |
|  | Asia/Oceania | 4 | Australia, China, Malaysia, Singapore |
|  | Europe | 11 | Czech Republic, Denmark, France, Germany, Italy, Latvia, Luxembourg, Moldova (Republic of), Portugal, Spain, Sweden |
| *Median Stringency index*<sup>1</sup> | Low Stringency (0-35) | 0 |  |
|  | Middle Stringency (36-65) | 12 | Australia, Czech Republic, Denmark, Germany, Latvia, Luxembourg, Mexico, Portugal, Singapore, Sweden, United States, Uruguay |
|  | High Stringency (66-100) | 13 | Argentina, Botswana, China, Colombia, France, Italy, Kenya, Malaysia, Moldova (Republic of), Nigeria, Panama, Spain, Uganda |
| *Number of Participants* | 200- 499 participants | 9 | Botswana, Italy, Kenya, Malaysia, Moldova (Republic of), Nigeria, Spain, Uganda, United States |
|  | 500 – 999 participants | 8 | Argentina, Australia, China, Germany, Luxembourg, Panama, Singapore, Uruguay |
|  | >999 participants | 8 | Colombia, Czech Republic, Denmark, France, Latvia, Mexico, Portugal, Sweden |

<sup>1</sup>Oxford Stringency Index: a measure to record the strictness of lockdown policies based on indicators such as school and workplace closure, and travel bans, rescaled to a value from 0 to 100 (100 = strictest). We measured the median during the period when the survey was open in each country.

**Supplemental Table 2.**
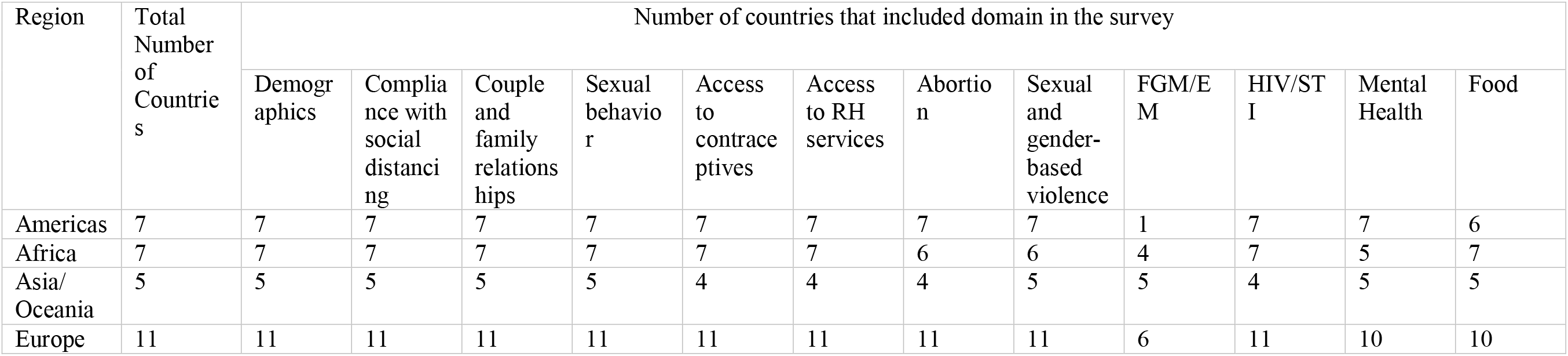
I-SHARE survey instrument components

| Region | Total Number of Countries | Number of countries that included domain in the survey |  |  |  |  |  |  |  |  |  |  |  |
| --- | --- | --- | --- | --- | --- | --- | --- | --- | --- | --- | --- | --- | --- |
|  |  | Demographics | Compliance with social distancing | Couple and family relationships | Sexual behavior | Access to contraceptives | Access to RH services | Abortion | Sexual and gender-based violence | FGM/EM | HIV/STI | Mental Health | Food |
| Americas | 7 | 7 | 7 | 7 | 7 | 7 | 7 | 7 | 7 | 1 | 7 | 7 | 6 |
| Africa | 7 | 7 | 7 | 7 | 7 | 7 | 7 | 6 | 6 | 4 | 7 | 5 | 7 |
| Asia/Oceania | 5 | 5 | 5 | 5 | 5 | 4 | 4 | 4 | 5 | 5 | 4 | 5 | 5 |
| Europe | 11 | 11 | 11 | 11 | 11 | 11 | 11 | 11 | 11 | 6 | 11 | 10 | 10 |

**Supplemental Table 3.**
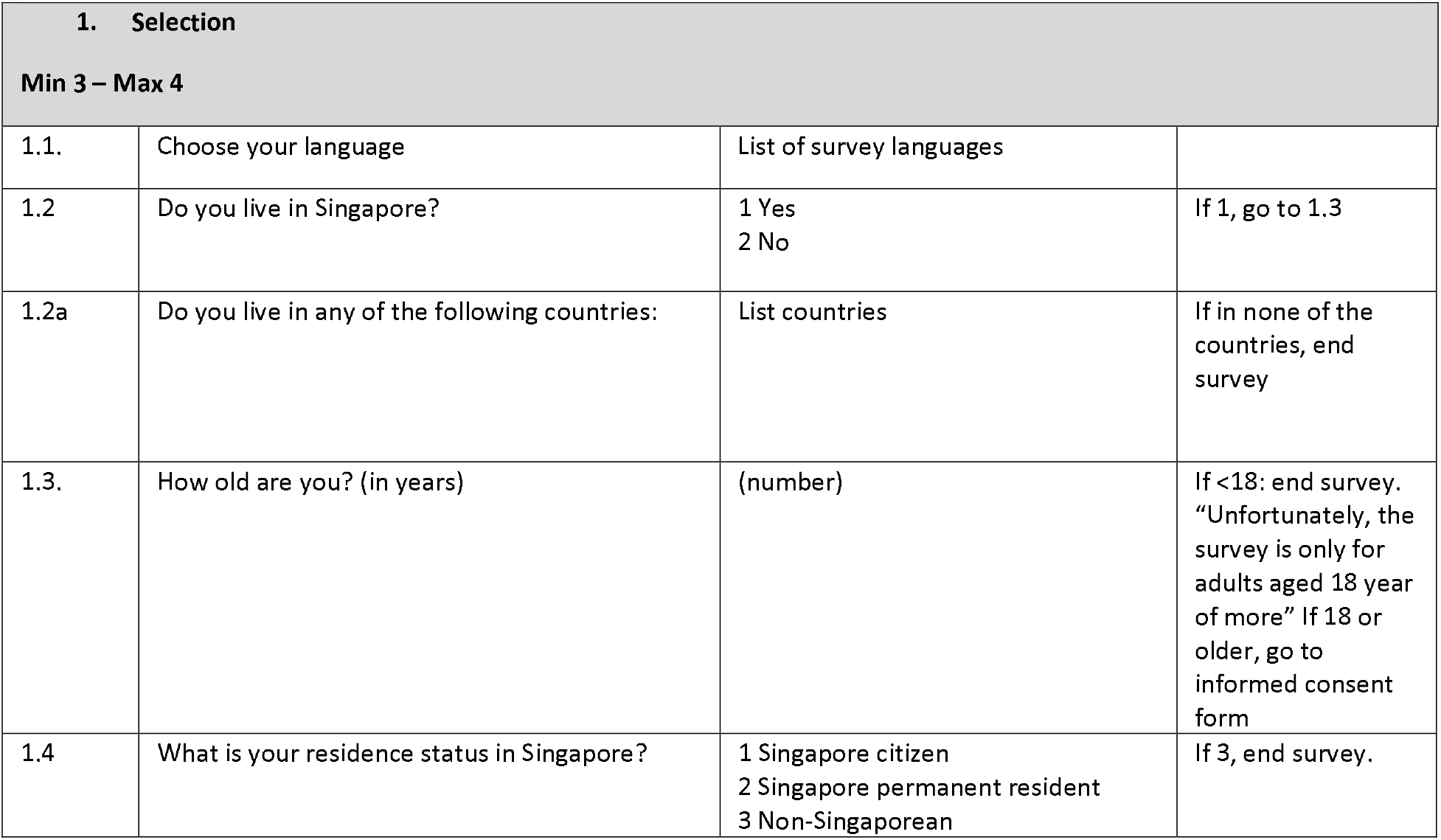
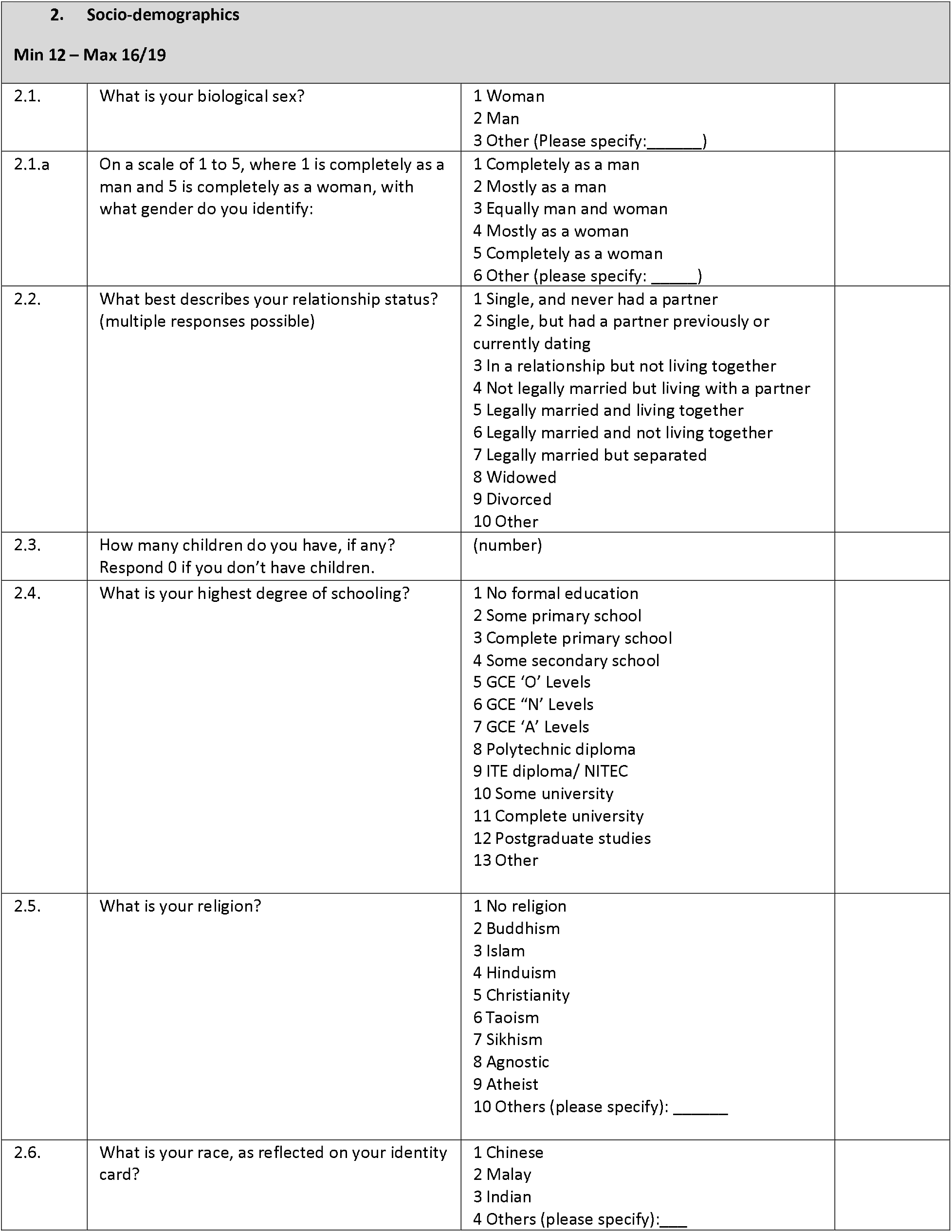
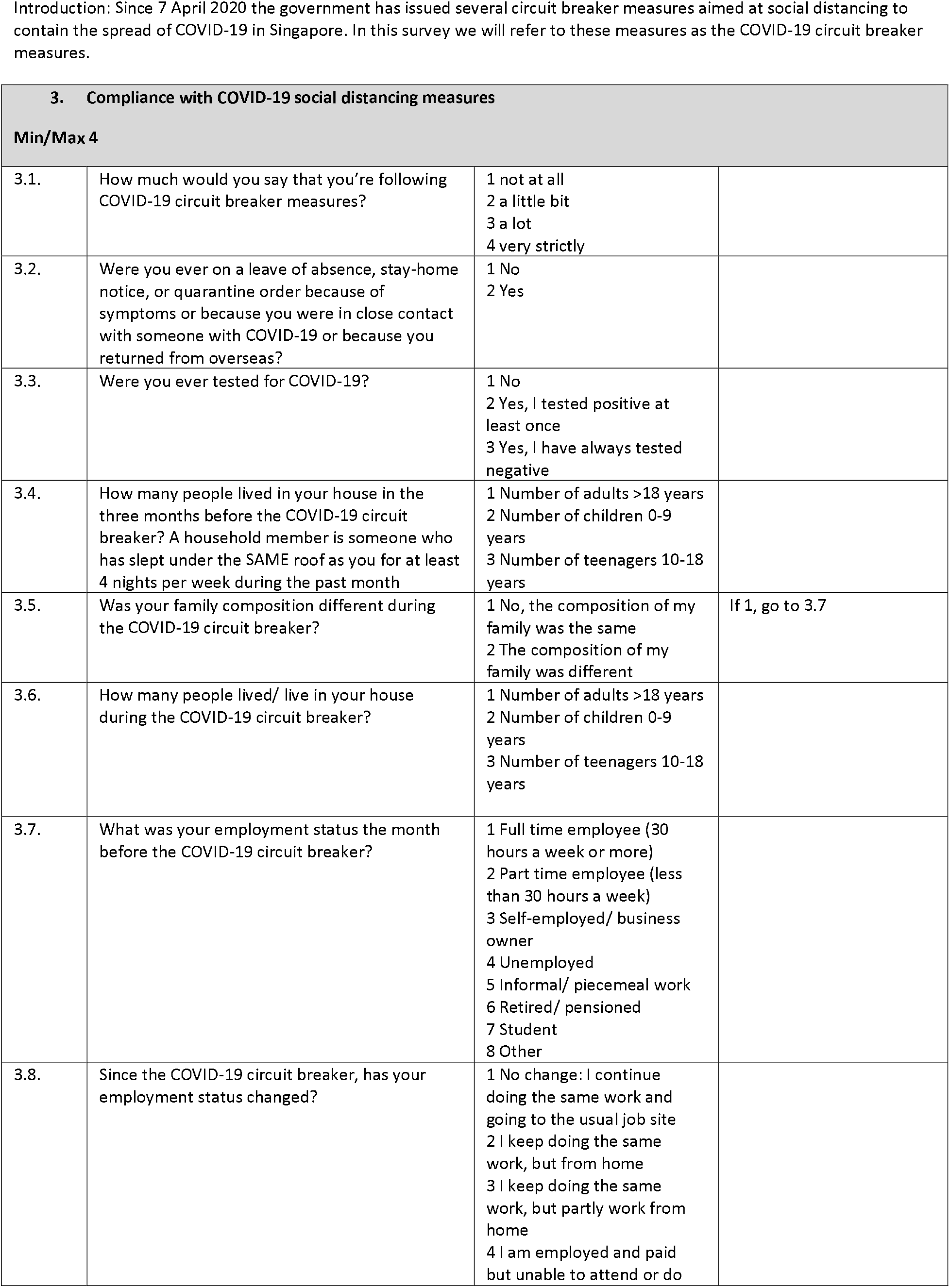
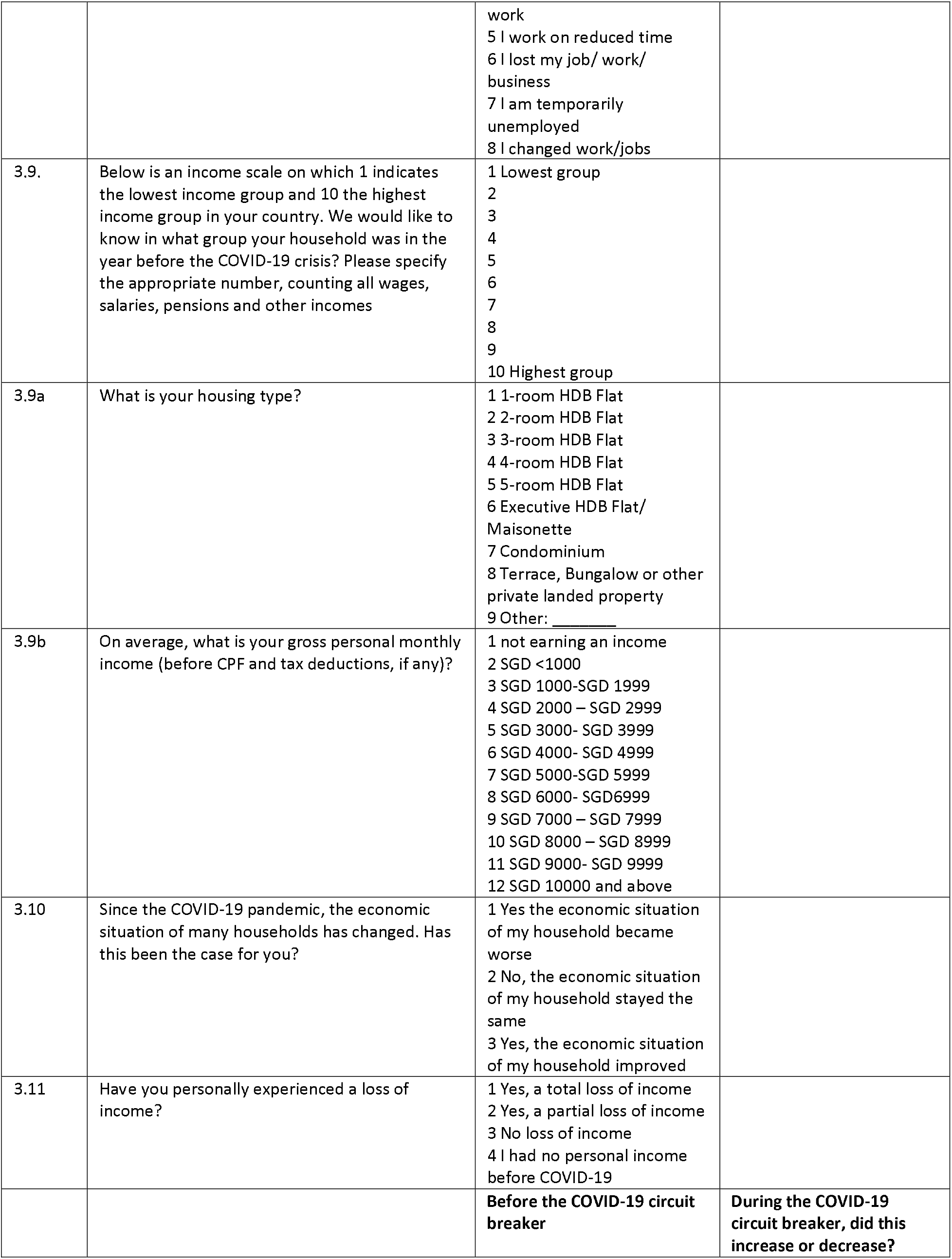
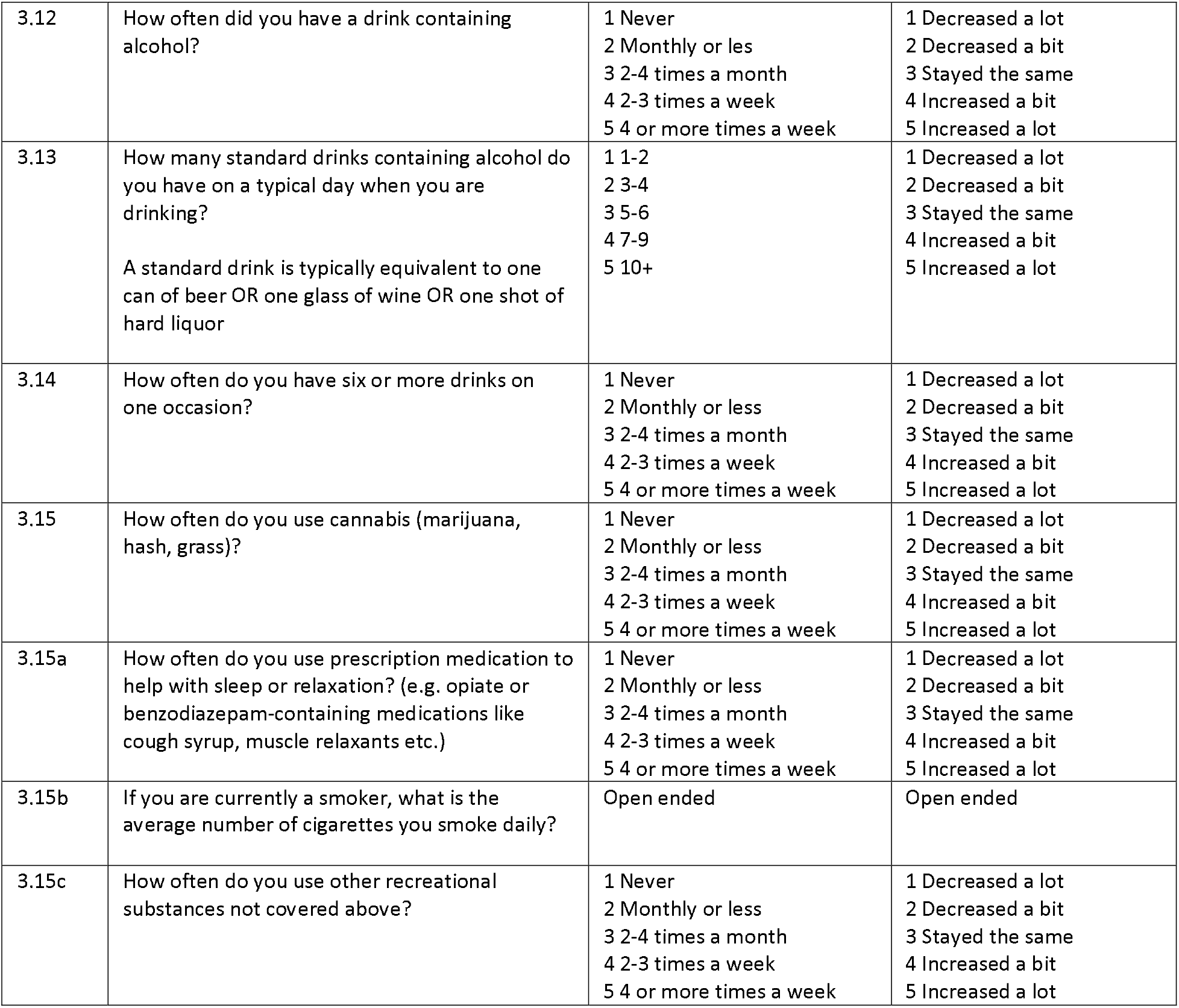
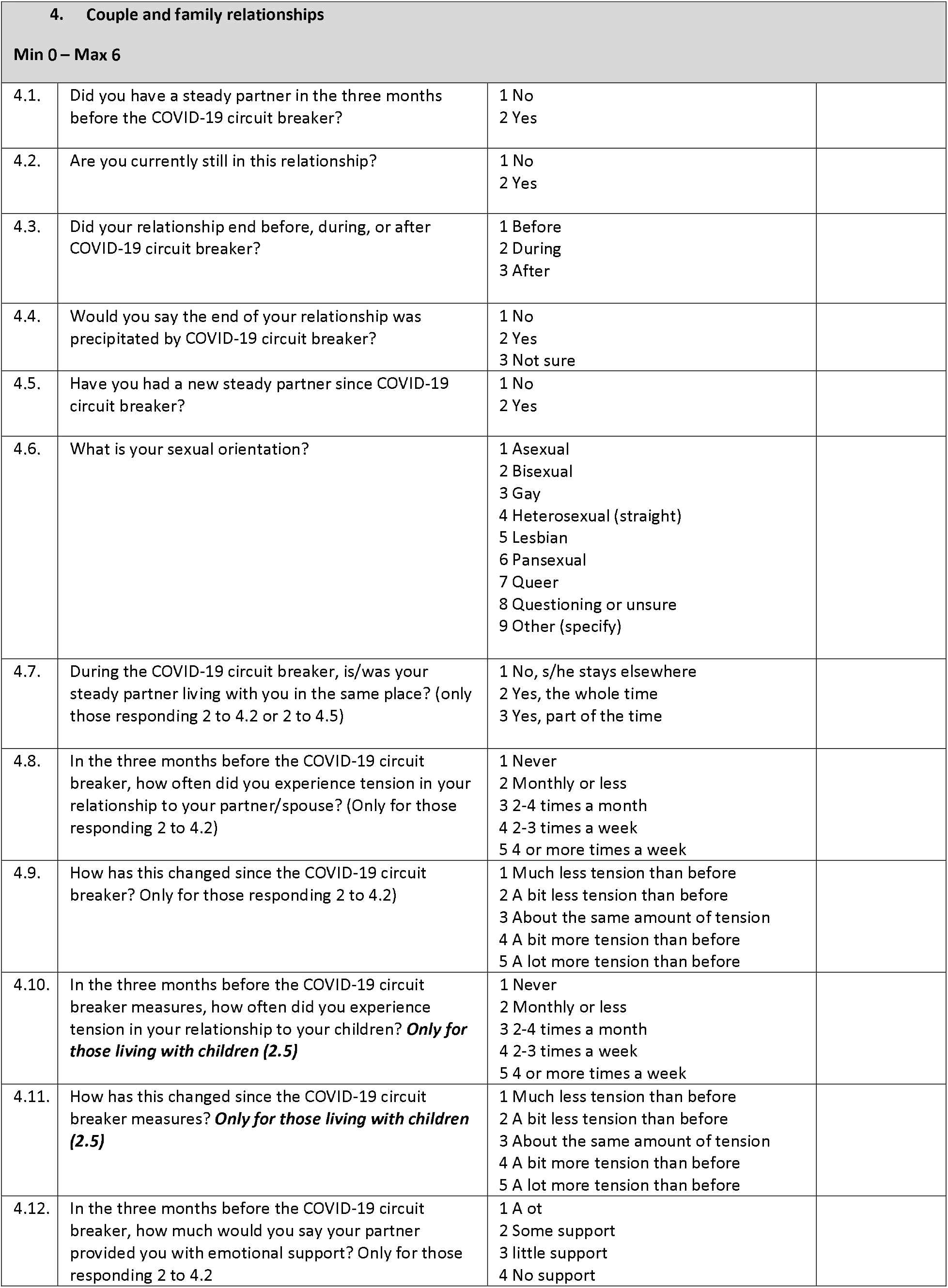
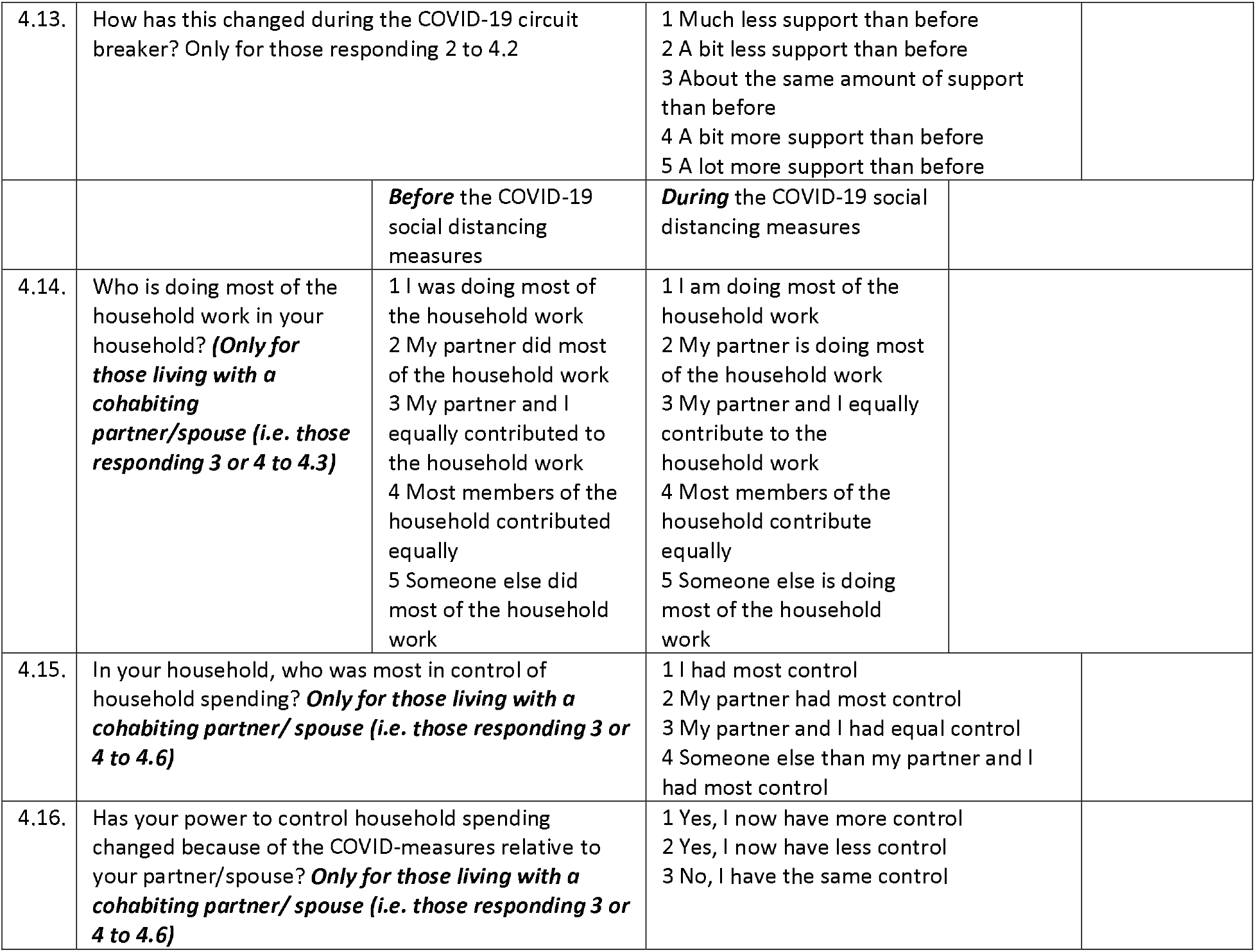
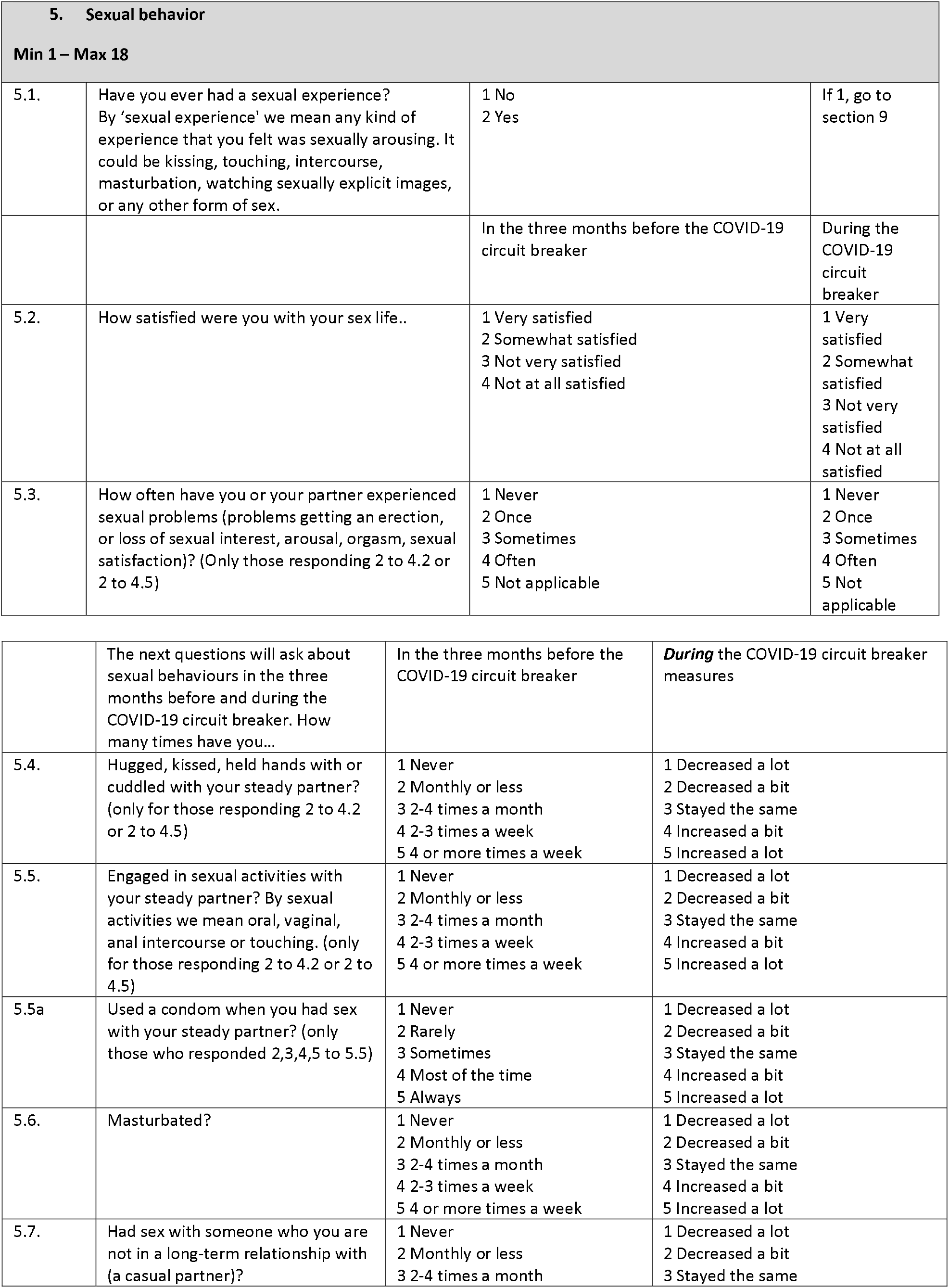
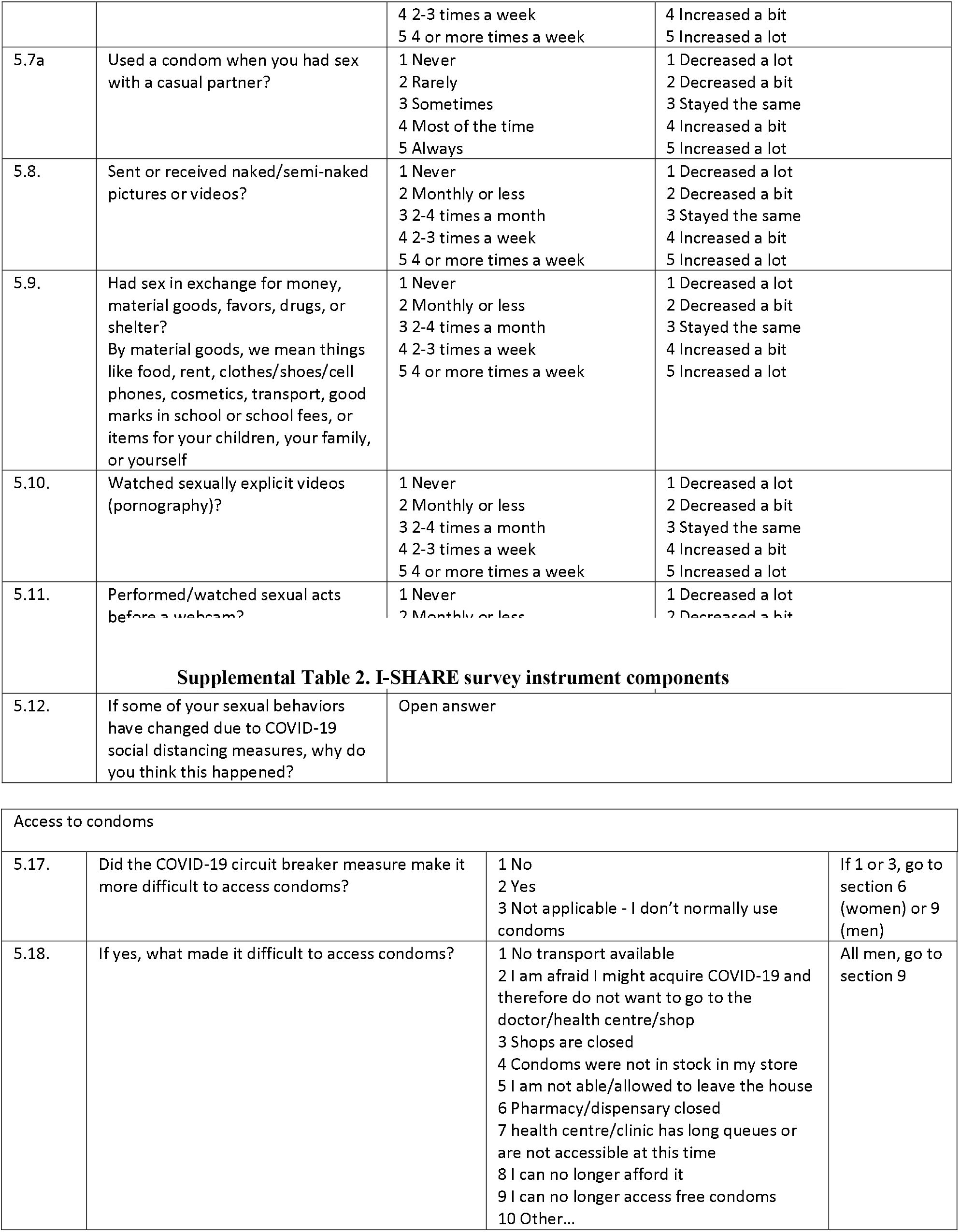
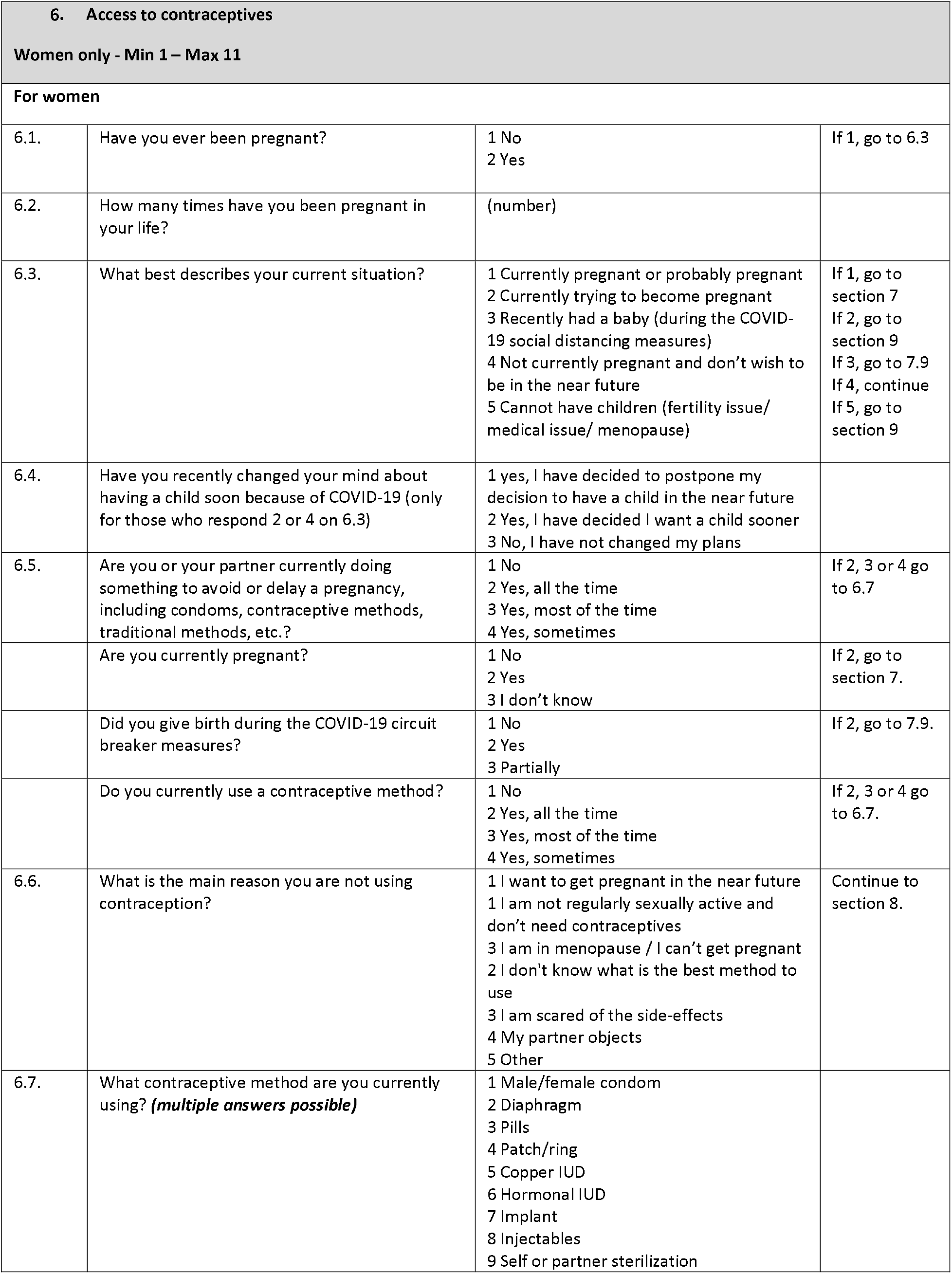
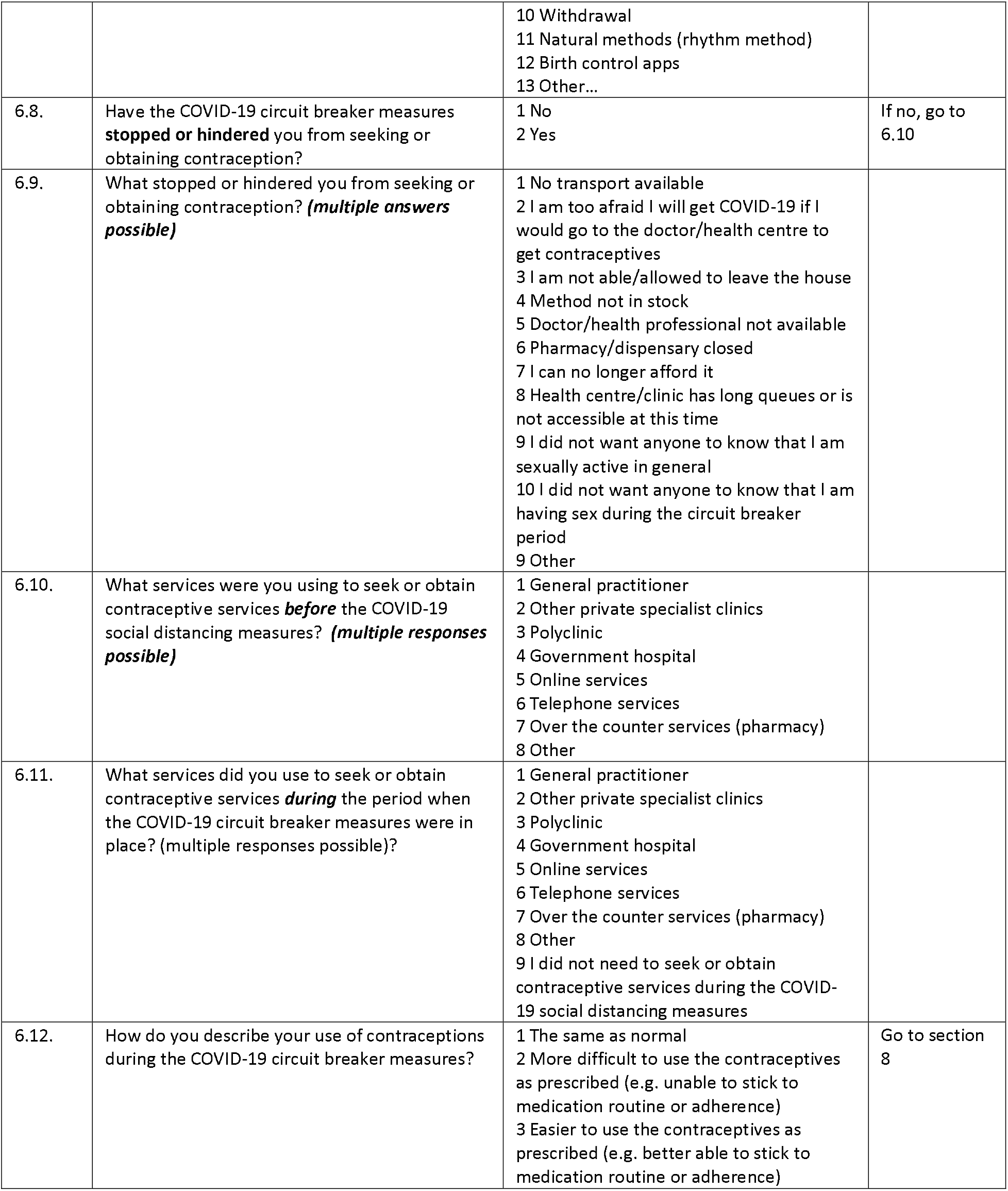
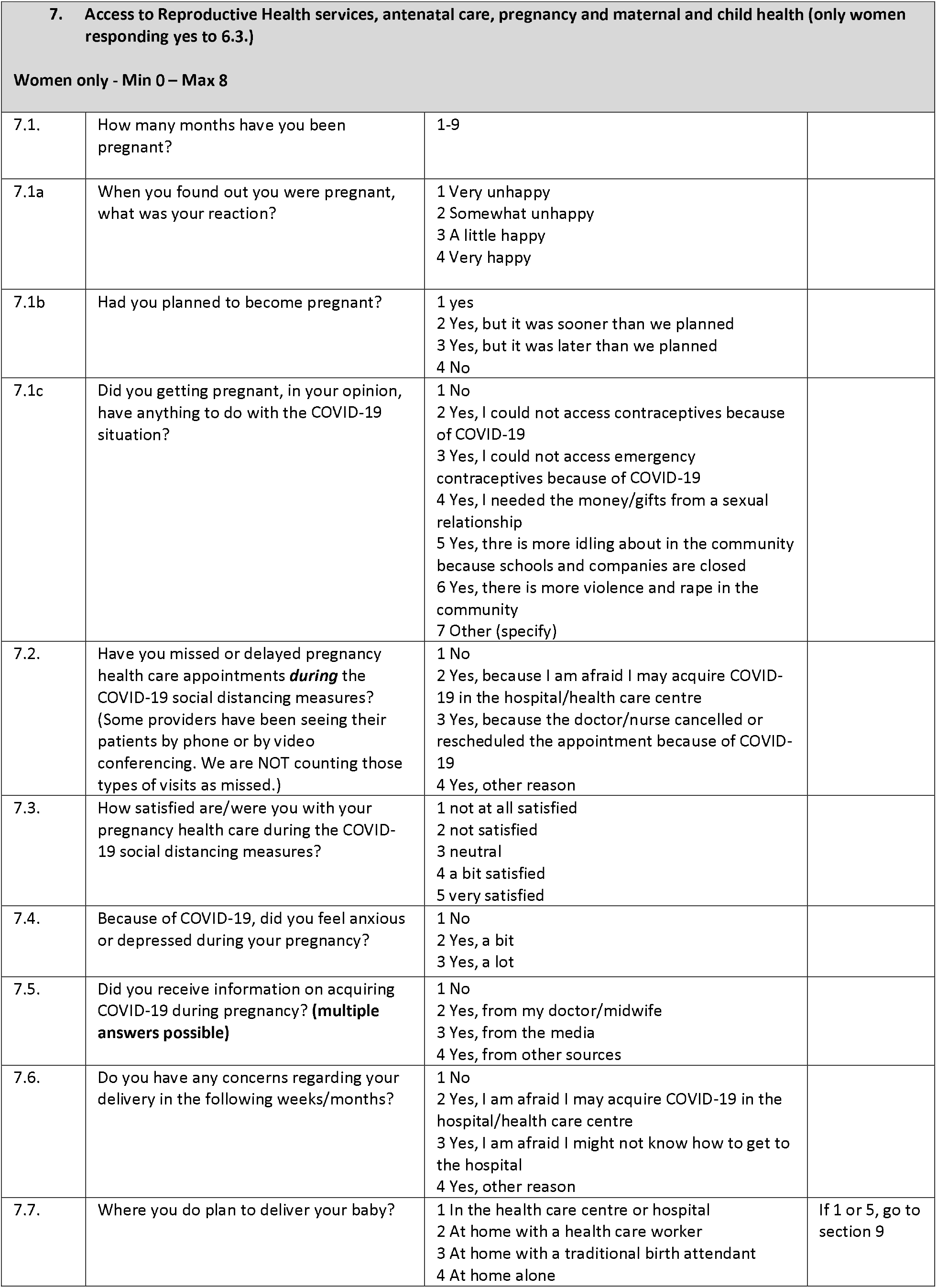
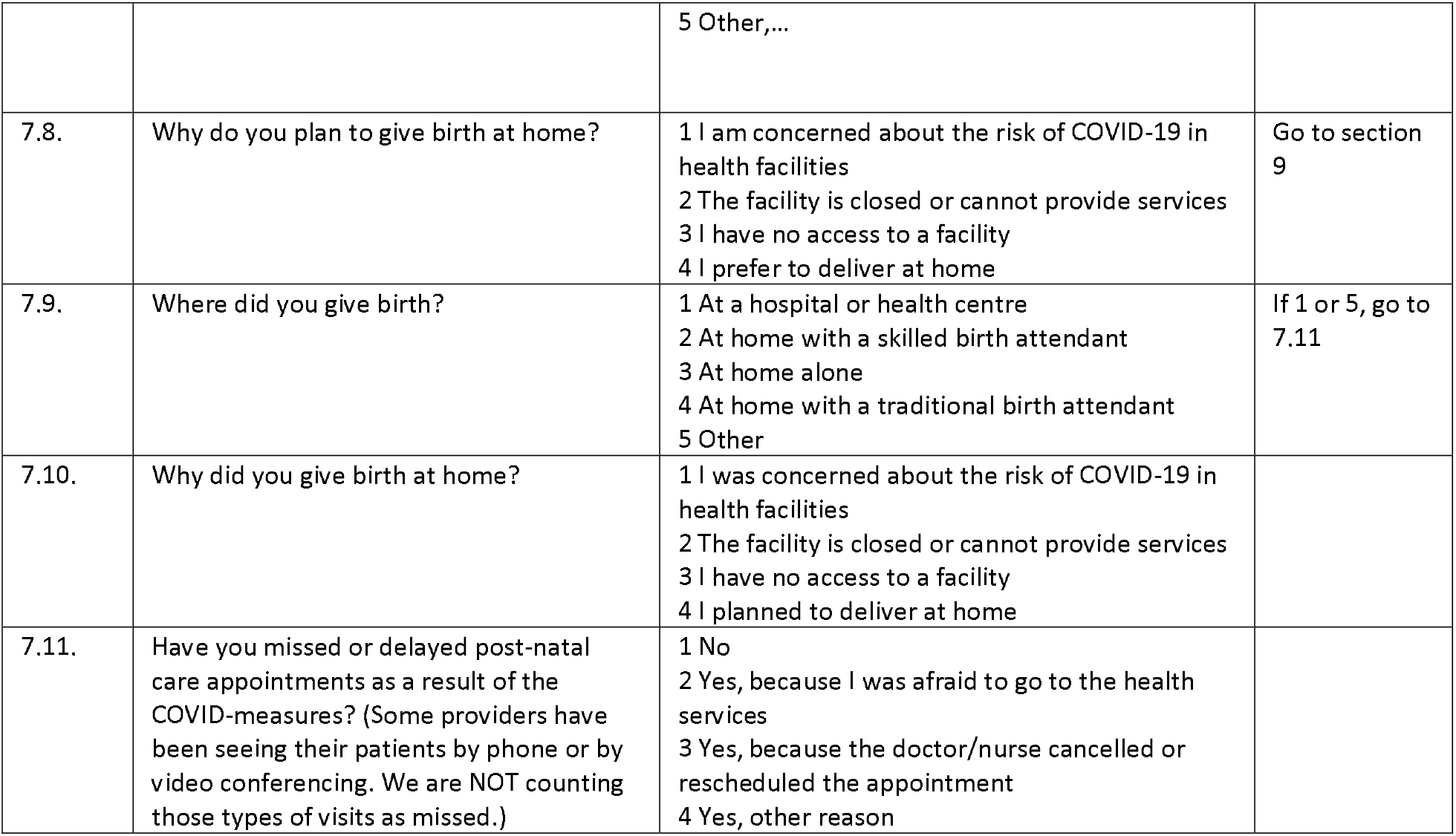
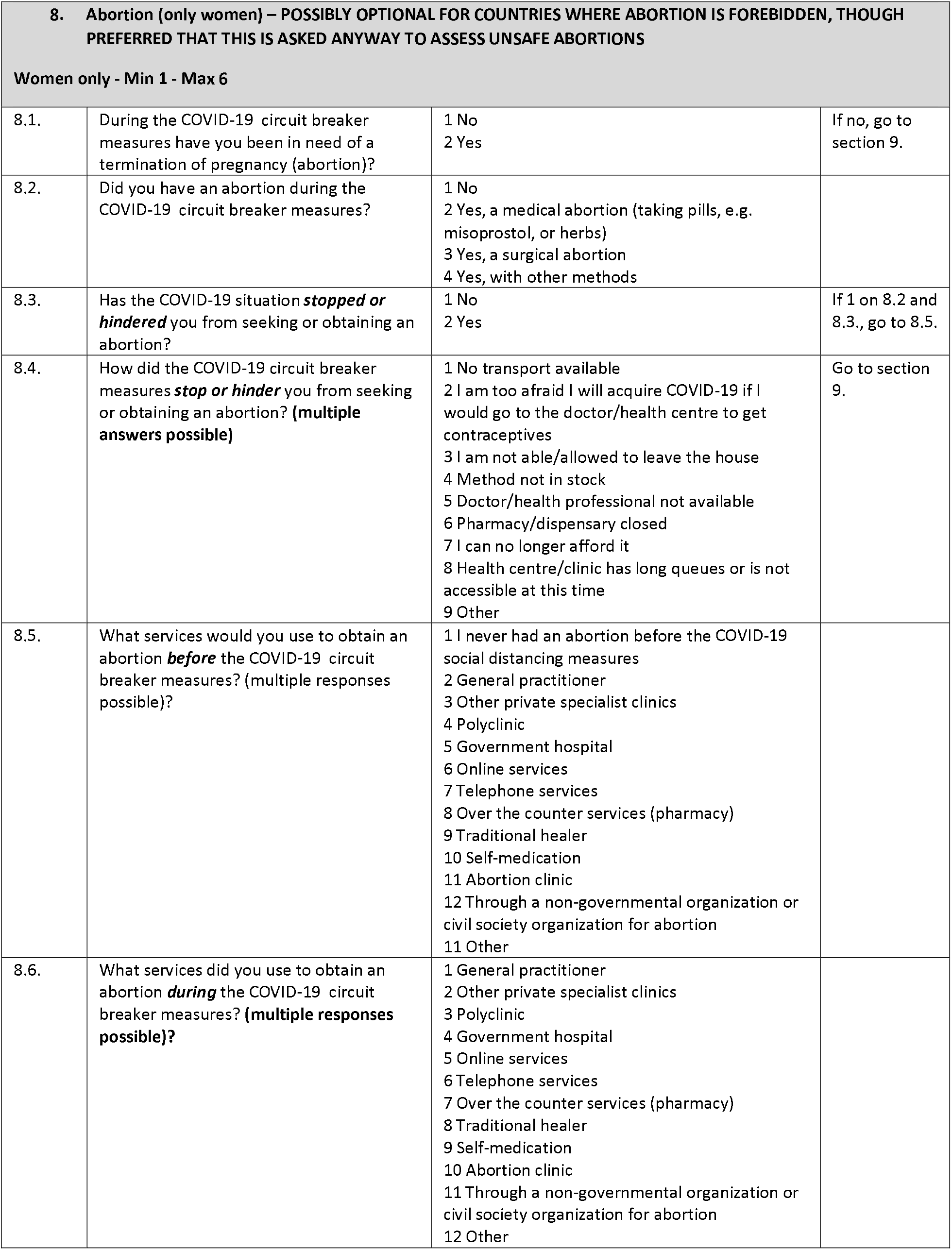
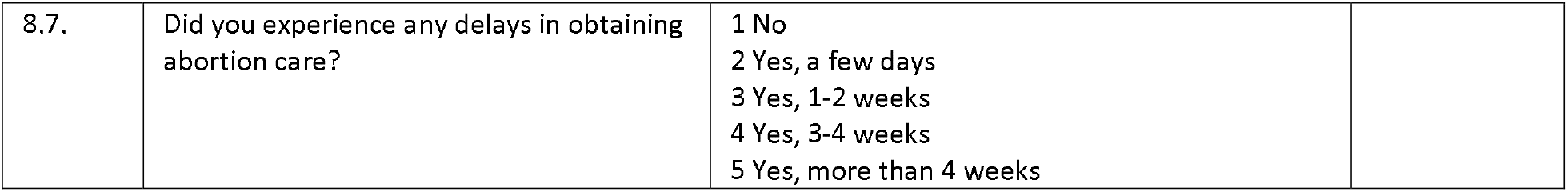
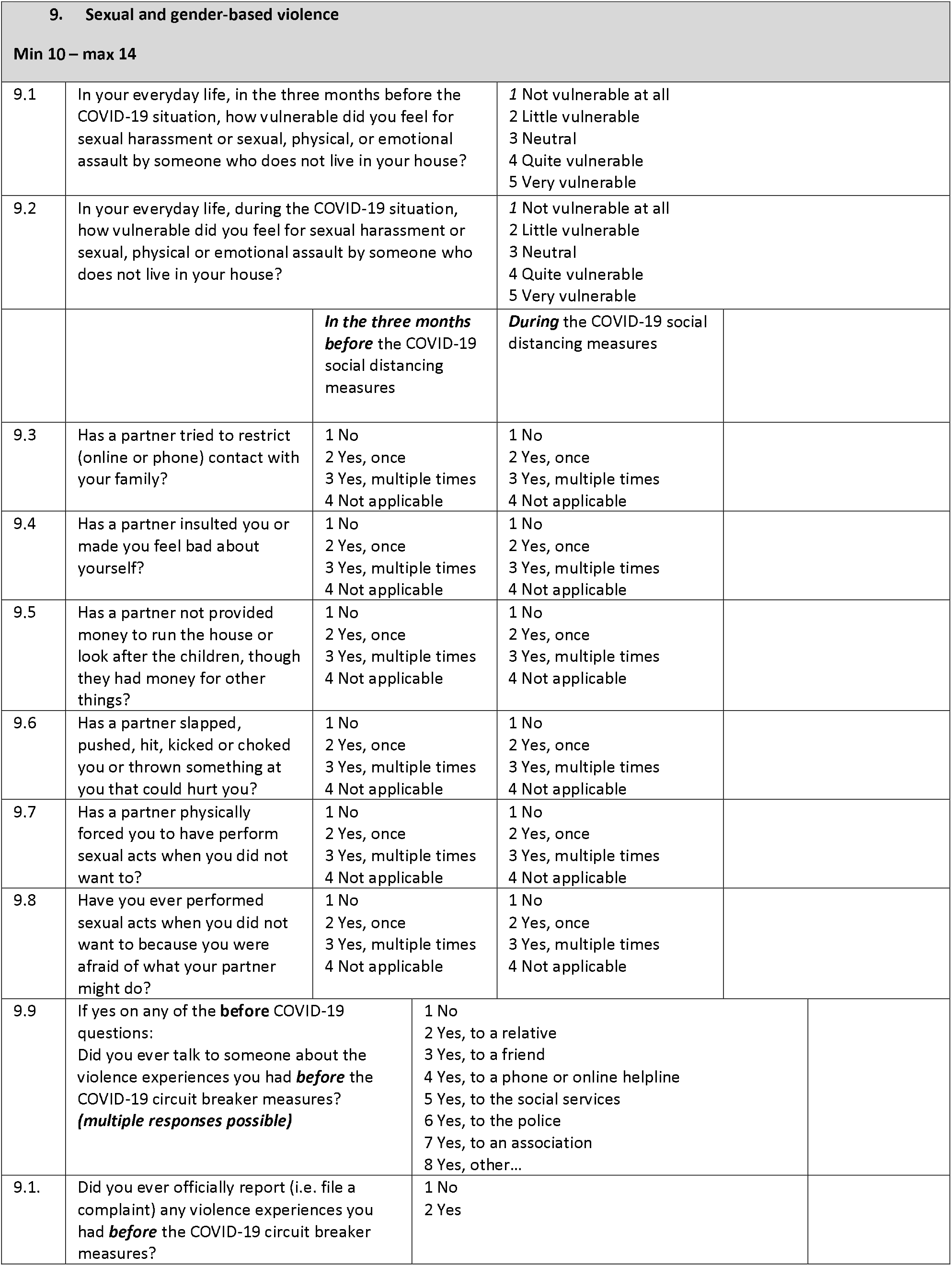
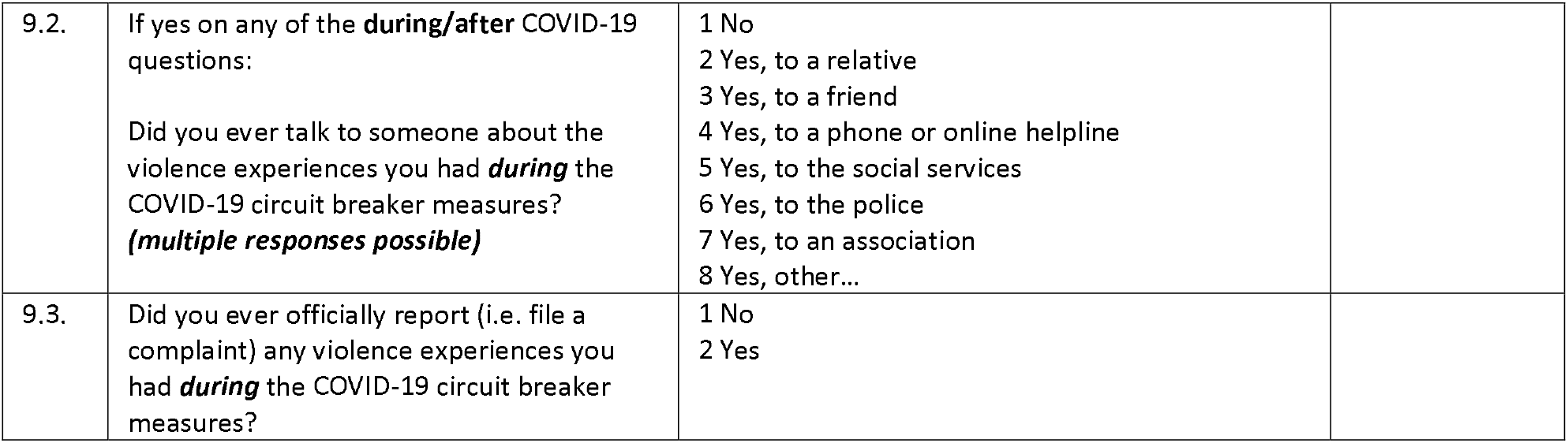
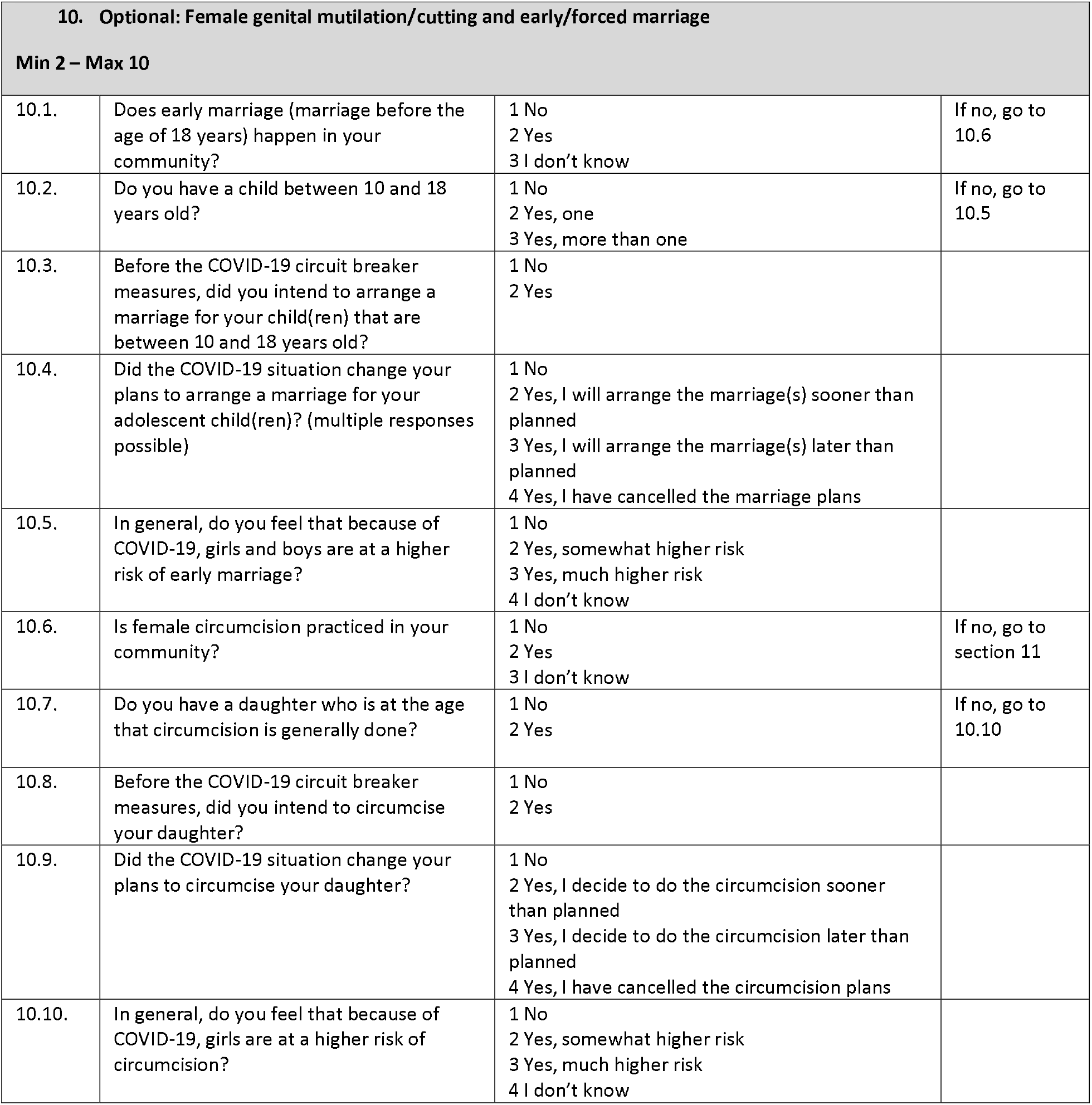
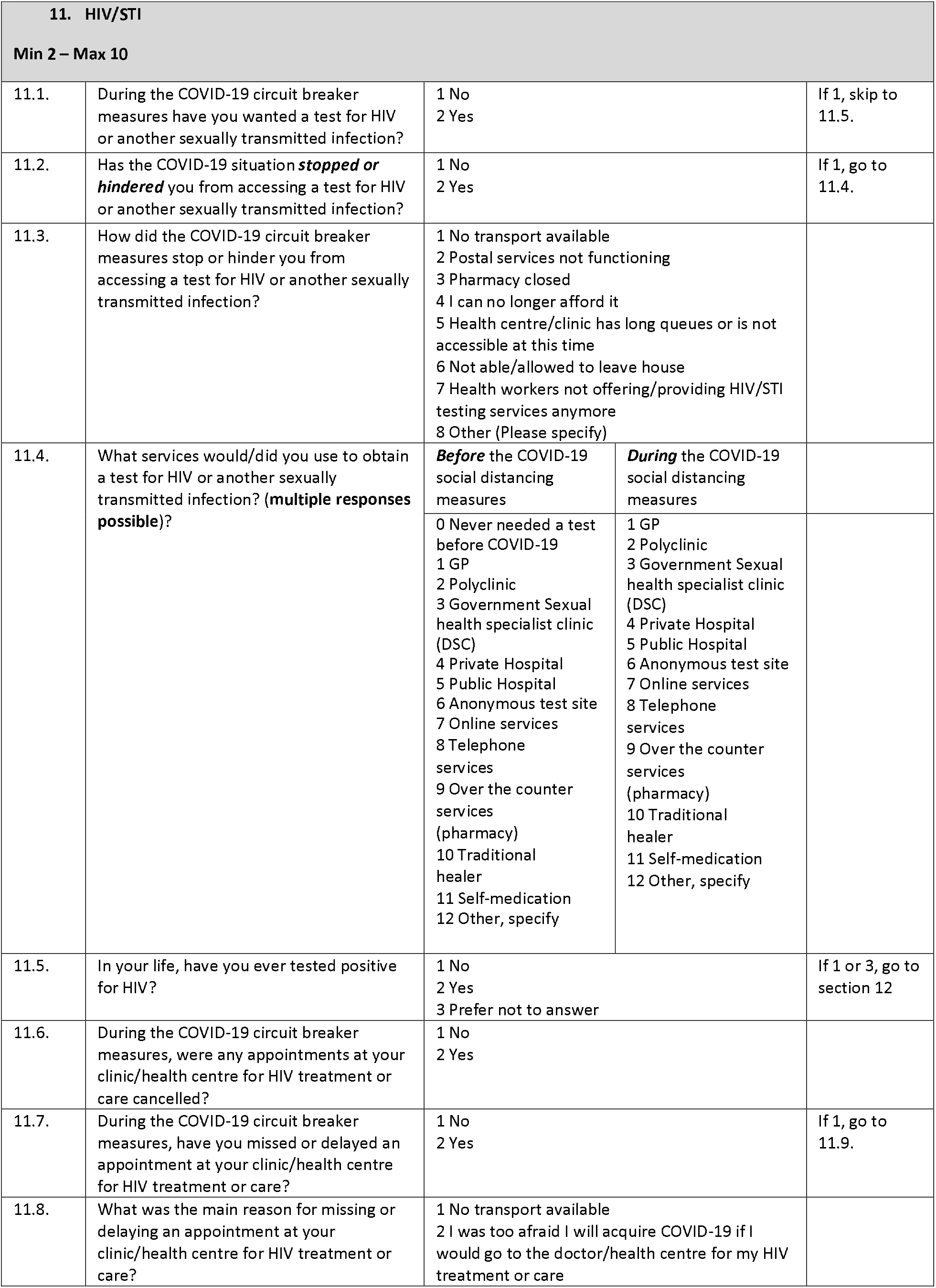
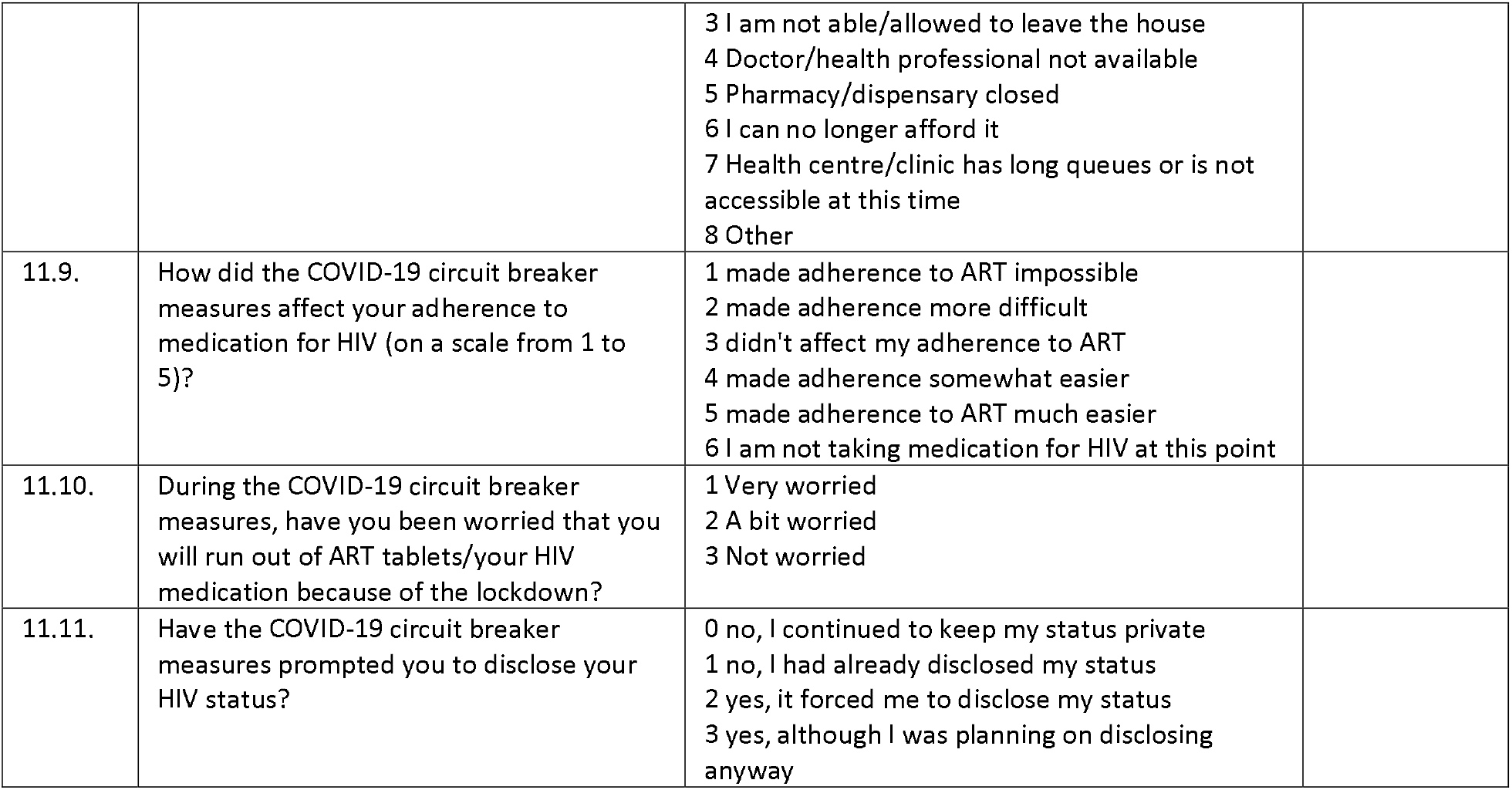
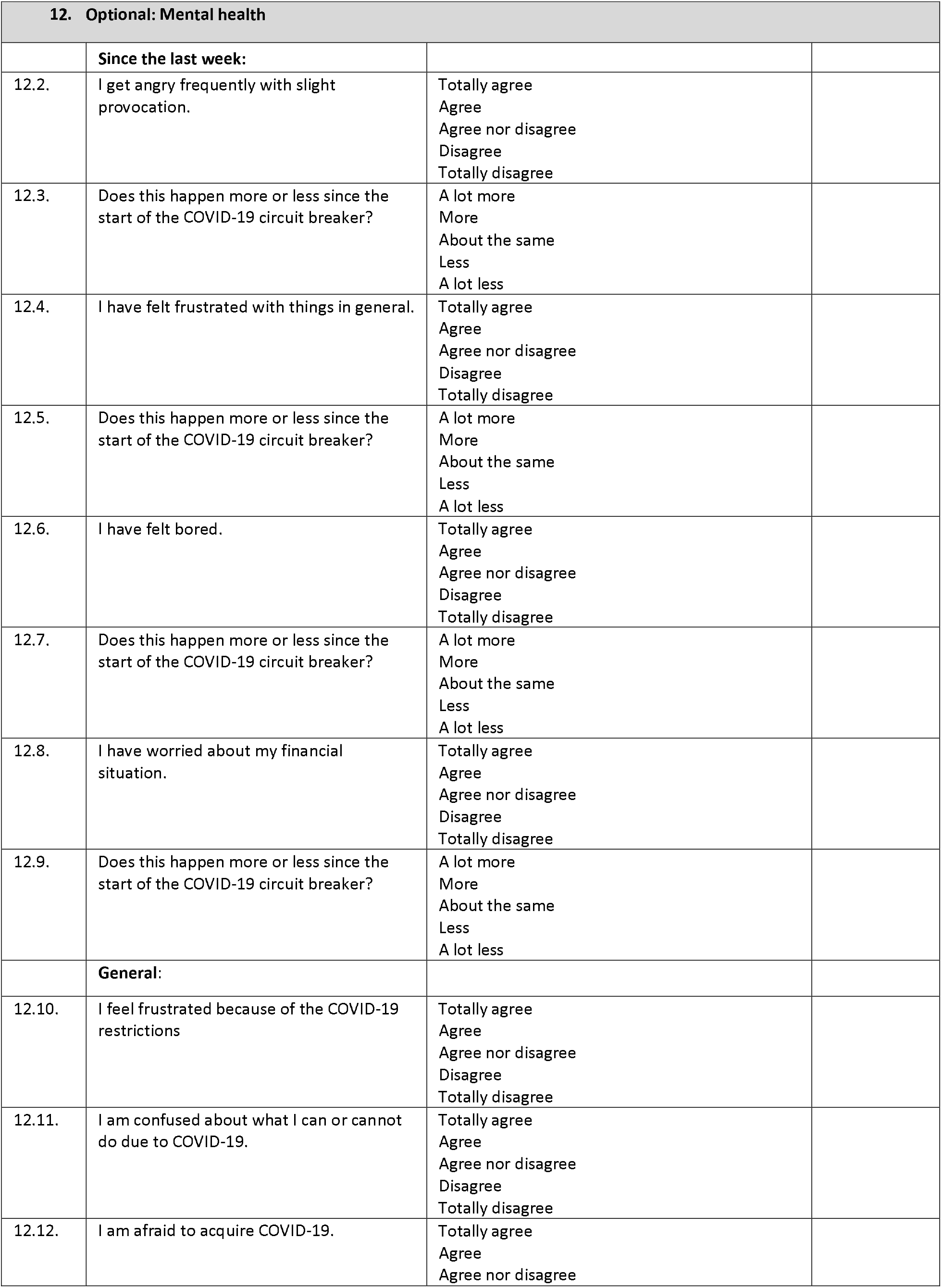
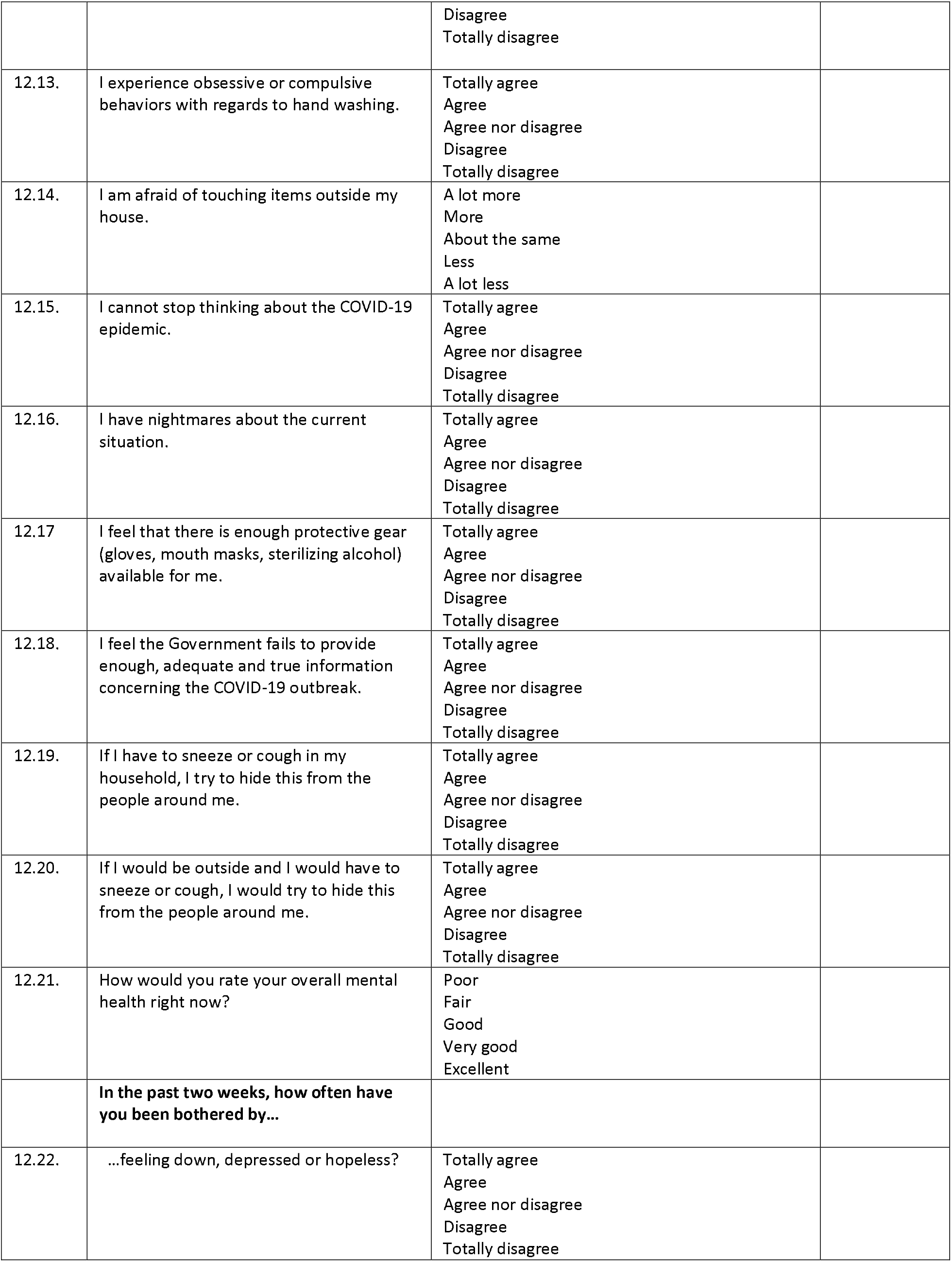
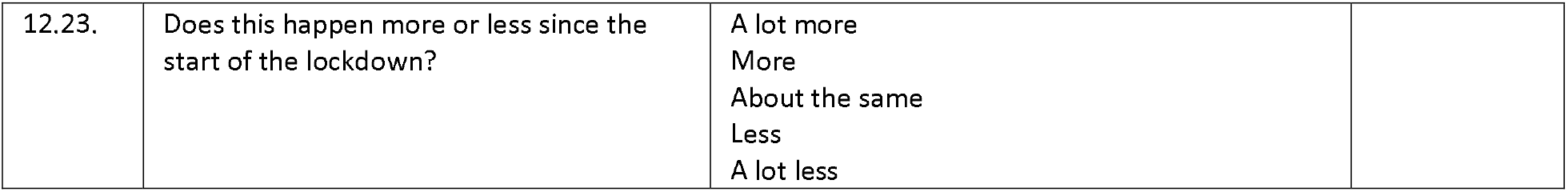
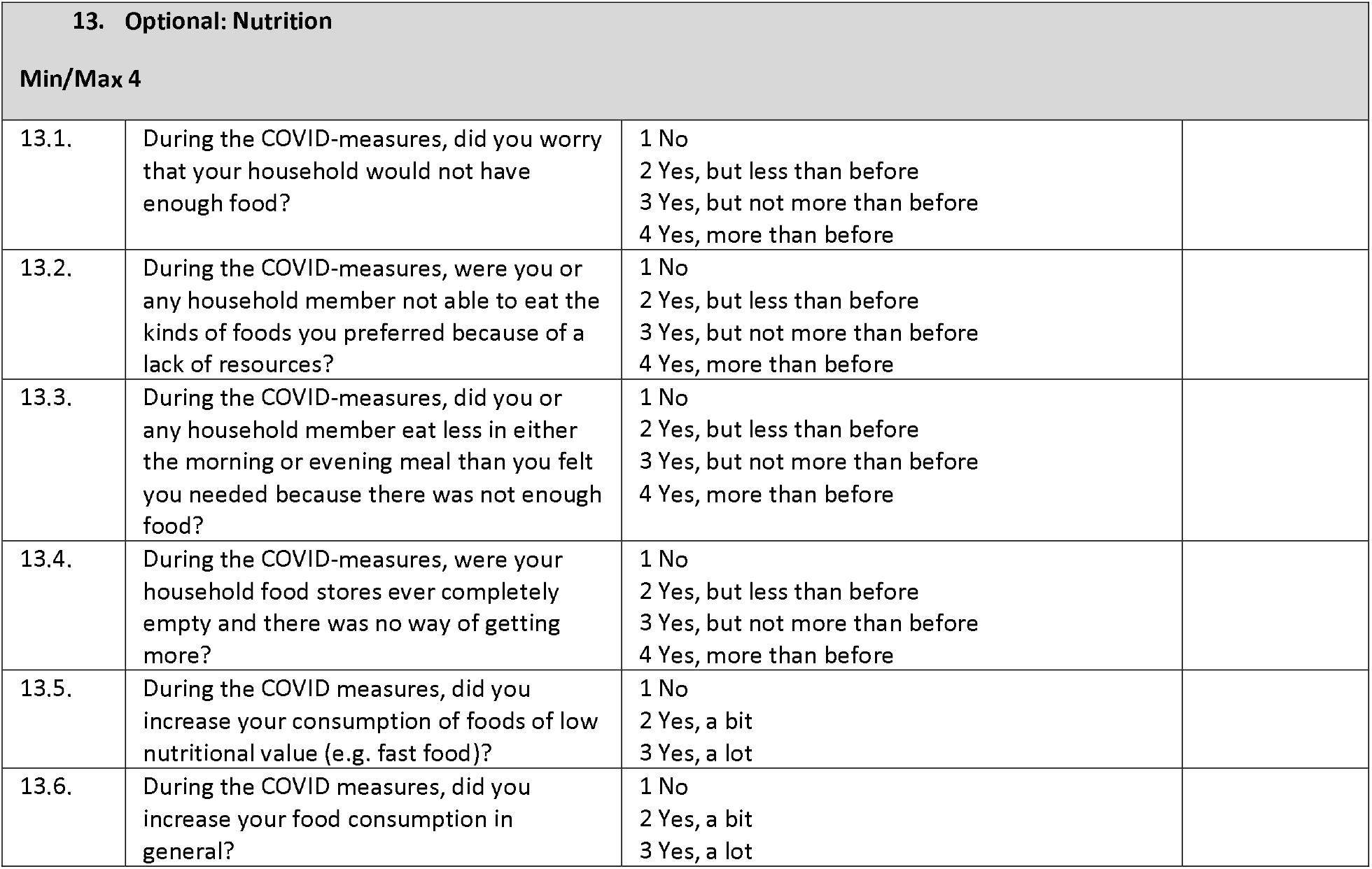
Survey instrument used in I-SHARE (Singapore Version)

**Supplemental Table 4.** COVID-19 variables in the I-SHARE multi-country survey, 2020-2021.

|  |  |  |
| --- | --- | --- |
| **How much would you say that you're following COVID-19 social distancing measures?** | **n** | **%** |
| Not at all | 316 | 1.4 |
| A little bit | 3558 | 15.7 |
| A lot | 13336 | 58.9 |
| Very strictly | 5432 | 24.0 |
| **Total** | **22642** | **100.0** |
| **Were you ever in (self-)isolation because of symptoms or because you were in close contact with someone with COVID-19 or because you returned from a country that had a large number of cases?** | **n** | **%** |
| No | 17327 | 76.6 |
| Yes | 5303 | 23.4 |
| **Total** | **22630** | **100.0** |
| **Were you ever tested for COVID-19?** |  |  |
| No | 14996 | 66.2 |
| Yes, I have always tested negative | 6421 | 28.4 |
| Yes, I have not received the results | 49 | 0.2 |
| Yes, I tested positive at least once | 1175 | 5.2 |
| **Total** | **22641** | **100.0** |
| **Since the COVID-pandemic, the economic situation of many households has changed. Has this been the case for you?** |  |  |
| Yes, the economic situation of my household became worse | 7235 | 32.0 |
| No, the economic situation of my household stayed the same | 14046 | 62.2 |
| Yes, the economic situation of my household became improved | 1302 | 5.8 |
| **Total** | **22583** | **100.0** |
| **Have you personally experienced a loss of income?** |  |  |
| Yes, a total loss of income | 1634 | 7.6 |
| Yes, a partial loss of income | 5495 | 25.5 |
| No loss of income | 11807 | 54.8 |
| I have no personal income before COVID-19 | 2607 | 12.1 |
| **Total** | **21543** | **100.0** |

**Supplemental Table 5.** AXIS Risk of Bias Assessment for I-SHARE country surveys (n=25)

**The Appraisal of the I-SHARE country specific surveys (1 of 3)**
| Components | Country |  |  |  |  |  |  |  |
| --- | --- | --- | --- | --- | --- | --- | --- | --- |
|  | Argentina | Australia | Botswana | China | Colombia | Czech Republic | Denmark | France |
| **Introduction** |  |  |  |  |  |  |  |  |
| 1. Were the aims/objectives of the study clear? | Yes | Yes | Yes | Yes | Yes | Yes | Yes | Yes |
| **Methods** |  |  |  |  |  |  |  |  |
| 2. Was the study design appropriate for the stated aim(s)? | Yes | Yes | Yes | Yes | Yes | Yes | Yes | Yes |
| 3. Was the sample size justified? | Yes | Yes | Yes | Yes | Yes | Yes | Yes | Yes |
| 4. Was the target/reference population clearly defined? (Is it clear who the research was about?) | Yes | Yes | Yes | Yes | Yes | Yes | Yes | Yes |
| 5. Was the sample frame taken from an appropriate population base so that it closely represented the target/reference population under investigation? | No | No | No | No | No | Yes | Yes | No |
| 6. Was the selection process likely to select subjects/participants that were representative of the target/reference population under investigation? | No | No | No | No | No | Yes | Yes | No |
| 7. Were measures undertaken to address and categorise non-responders? | No | No | No | No | No | Yes | Yes | No |
| 8. Were the risk factor and outcome variables measured appropriate to the aims of the study? | Yes | Yes | Yes | Yes | Yes | Yes | Yes | Yes |
| 9. Were the risk factor and outcome variables measured correctly using instruments/measurements that had been trialled, piloted or published previously? | Yes | Yes | Yes | Yes | Yes | Yes | Yes | Yes |
| 10. Is it clear what was used to determined statistical significance and/or precision estimates? (e.g., p-values, CIs) | Yes | Yes | Yes | Yes | Yes | Yes | Yes | Yes |
| 11. Were the methods (including statistical methods) sufficiently described to enable them to be repeated? | Yes | Yes | Yes | Yes | Yes | Yes | Yes | Yes |
| **Results** |  |  |  |  |  |  |  |  |
| 12. Were the basic data adequately described? | Yes | Yes | Yes | Yes | Yes | Yes | Yes | Yes |
| 13. Does the response rate raise concerns about non-response bias? | DNK | DNK | DNK | DNK | DNK | Yes | Yes | DNK |
| 14. If appropriate, was information about non-responders described? | NA | NA | NA | NA | NA | No | No | NA |
| 15. Were the results internally consistent? | Yes | Yes | Yes | Yes | Yes | Yes | Yes | Yes |
| 16. Were the results for the analyses described in the methods, presented? | Yes | Yes | Yes | Yes | Yes | Yes | Yes | Yes |

**The Appraisal of the I-SHARE country specific surveys (2 of 3)**
| **Components** | Germany | Italy | Kenya | Latvia | Luxembourg | Malaysia | Mexico | Moldova |
| --- | --- | --- | --- | --- | --- | --- | --- | --- |
| **Introduction** |  |  |  |  |  |  |  |  |
| 1. Were the aims/objectives of the study clear? | Yes | Yes | Yes | Yes | Yes | Yes | Yes | Yes |
| **Methods** |  |  |  |  |  |  |  |  |
| 2. Was the study design appropriate for the stated aim(s)? | Yes | Yes | Yes | Yes | Yes | Yes | Yes | Yes |
| 3. Was the sample size justified? | Yes | Yes | Yes | Yes | Yes | Yes | Yes | Yes |
| 4. Was the target/reference population clearly defined? (Is it clear who the research was about?) | Yes | Yes | Yes | Yes | Yes | Yes | Yes | Yes |
| 5. Was the sample frame taken from an appropriate population base so that it closely represented the target/reference population under investigation? | No | No | No | No | No | No | No | No |
| 6. Was the selection process likely to select subjects/participants that were representative of the target/reference population under investigation? | No | No | No | No | No | No | No | No |
| 7. Were measures undertaken to address and categorize non-responders? | No | No | No | No | No | No | No | No |
| 8. Were the risk factor and outcome variables measured appropriate to the aims of the study? | Yes | Yes | Yes | Yes | Yes | Yes | Yes | Yes |
| 9. Were the risk factor and outcome variables measured correctly using instruments/ measurements that had been trialled, piloted, or published previously? | Yes | Yes | Yes | Yes | Yes | Yes | Yes | Yes |
| 10. Is it clear what was used to determined statistical significance and/or precision estimates? (eg, p values, CIs) | Yes | Yes | Yes | Yes | Yes | Yes | Yes | Yes |
| 11. Were the methods (including statistical methods) sufficiently described to enable them to be repeated? | Yes | Yes | Yes | Yes | Yes | Yes | Yes | Yes |
| **Results** |  |  |  |  |  |  |  |  |
| 12. Were the basic data adequately described? | Yes | Yes | Yes | Yes | Yes | Yes | Yes | Yes |
| 13. Does the response rate raise concerns about non-response bias? | DNK | DNK | DNK | DNK | DNK | DNK | DNK | DNK |
| 14. If appropriate, was information about non-responders described? | NA | NA | NA | NA | NA | NA | NA | NA |
| 15. Were the results internally consistent? | Yes | Yes | Yes | Yes | Yes | Yes | Yes | Yes |
| 16. Were the results for the analyses described in the methods, presented? | Yes | Yes | Yes | Yes | Yes | Yes | Yes | Yes |

**The Appraisal of the I-SHARE country specific surveys (3 of 3)**
| **Components** | Nigeria | Panama | Portugal | Singapore | Spain | Sweden | Uganda | Uruguay | USA |
| --- | --- | --- | --- | --- | --- | --- | --- | --- | --- |
| **Introduction** |  |  |  |  |  |  |  |  |  |
| 1. Were the aims/objectives of the study clear? | Yes | Yes | Yes | Yes | Yes | Yes | Yes | Yes | Yes |
| **Methods** |  |  |  |  |  |  |  |  |  |
| 2. Was the study design appropriate for the stated aim(s)? | Yes | Yes | Yes | Yes | Yes | Yes | Yes | Yes | Yes |
| 3. Was the sample size justified? | Yes | Yes | Yes | Yes | Yes | Yes | Yes | Yes | Yes |
| 4. Was the target/reference population clearly defined? (Is it clear who the research was about?) | Yes | Yes | Yes | Yes | Yes | Yes | Yes | Yes | Yes |
| 5. Was the sample frame taken from an appropriate population base so that it closely represented the target/reference population under investigation? | No | No | No | No | No | No | No | No | No |
| 6. Was the selection process likely to select subjects/participants that were representative of the target/reference population under investigation? | No | No | No | No | No | No | No | No | No |
| 7. Were measures undertaken to address and categorise non-responders? | No | No | No | No | No | No | No | No | No |
| 8. Were the risk factor and outcome variables measured appropriate to the aims of the study? | Yes | Yes | Yes | Yes | Yes | Yes | Yes | Yes | Yes |
| 9. Were the risk factor and outcome variables measured correctly using instruments/measurements that had been trialled, piloted, or published previously? | Yes | Yes | Yes | Yes | Yes | Yes | Yes | Yes | Yes |
| 10. Is it clear what was used to determined statistical significance and/or precision estimates? (e.g., p-values, CIs) | Yes | Yes | Yes | Yes | Yes | Yes | Yes | Yes | Yes |
| 11. Were the methods (including statistical methods) sufficiently described to enable them to be repeated? | Yes | Yes | Yes | Yes | Yes | Yes | Yes | Yes | Yes |
| **Results** |  |  |  |  |  |  |  |  |  |
| 12. Were the basic data adequately described? | Yes | Yes | Yes | Yes | Yes | Yes | Yes | Yes | Yes |
| 13. Does the response rate raise concerns about non-response bias? | DNK | DNK | DNK | DNK | DNK | DNK | DNK | DNK | DNK |
| 14. If appropriate, was information about non-responders described? | NA | NA | NA | NA | NA | NA | NA | NA | NA |
| 15. Were the results internally consistent? | Yes | Yes | Yes | Yes | Yes | Yes | Yes | Yes | Yes |
| 16. Were the results for the analyses described in the methods, presented? | Yes | Yes | Yes | Yes | Yes | Yes | Yes | Yes | Yes |

DNK (Do not know), NA (Not Applicable)

**Supplemental Table 7.** GRADE Framework to assess quality of evidence for meta-analysis outcomes

| **Outcomes** | **Effect Estimate (95%CI)** | **Certainty in Effect Estimates** | **Notes on Quality** |
| --- | --- | --- | --- |
| Hindered access to HIV/STI testing during COVID-19 measures | 32.3% (23.9 – 42.1%) | Moderate | Observational studies start at low quality. While risk of bias due to largely convenience sampling may further negatively impact certainty, quality of evidence was rated upwards due to large magnitude of effect, and a large sample size. |
| Experiencing physical and sexual violence during COVID-19 measures | 4.4% (3.4 – 5.4%) | Moderate | Observational studies start at low quality. While risk of bias due to largely convenience sampling may further negatively impact certainty, quality of evidence was rated upwards due to a large sample size |
| Decreased condom use with casual partners during COVID-19 measures | 5.8% (5.4 – 8.2%) | Moderate | Observational studies start at low quality. While risk of bias due to largely convenience sampling may further negatively impact certainty, quality of evidence was rated upwards due to a large sample size |

---

**Supplemental Figure 1.**
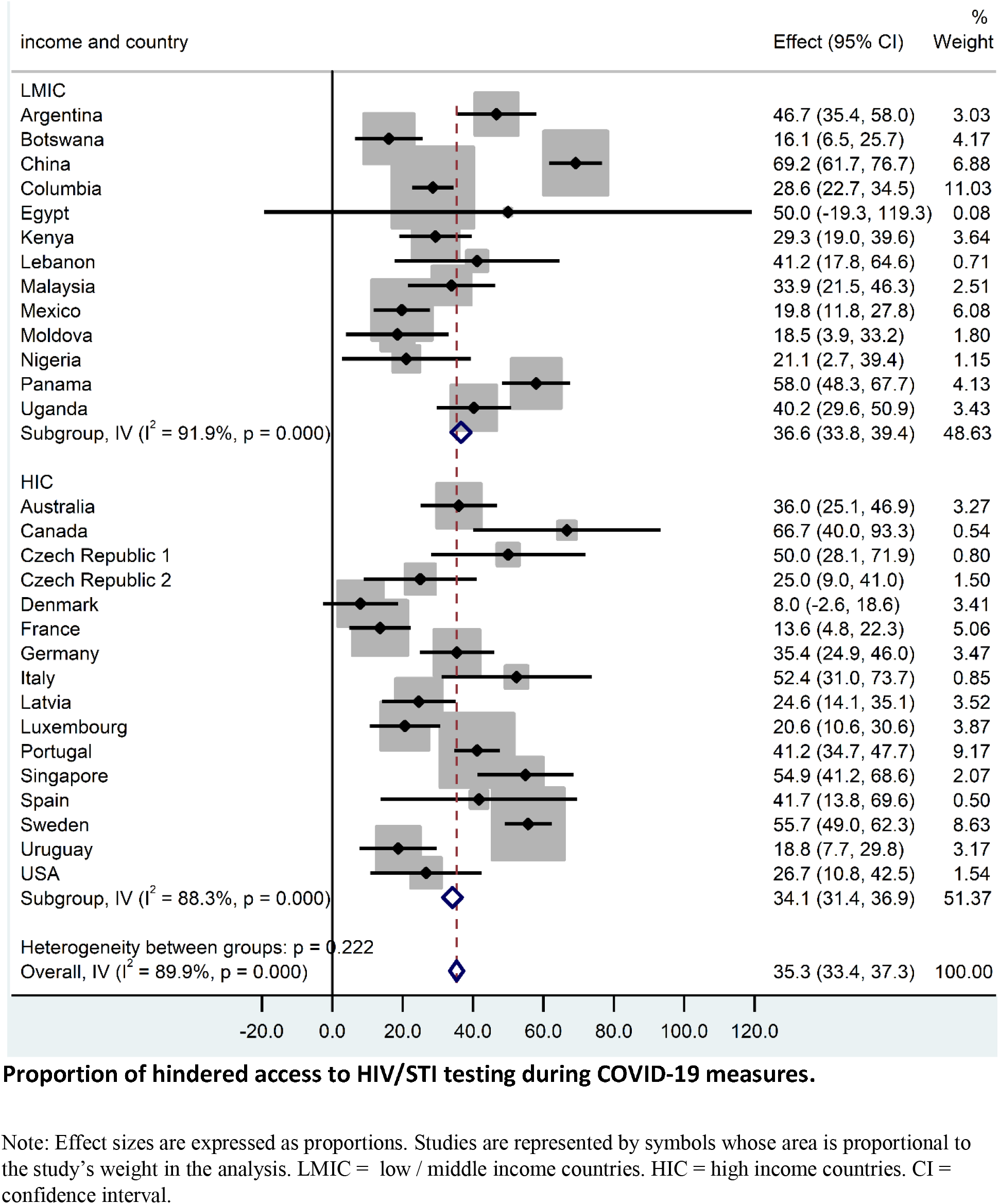
Forest plot for random-effects meta-analysis of the association between country income level and access to HIV/STI testing during COVID-19 measures.

**Supplemental Figure 2.**
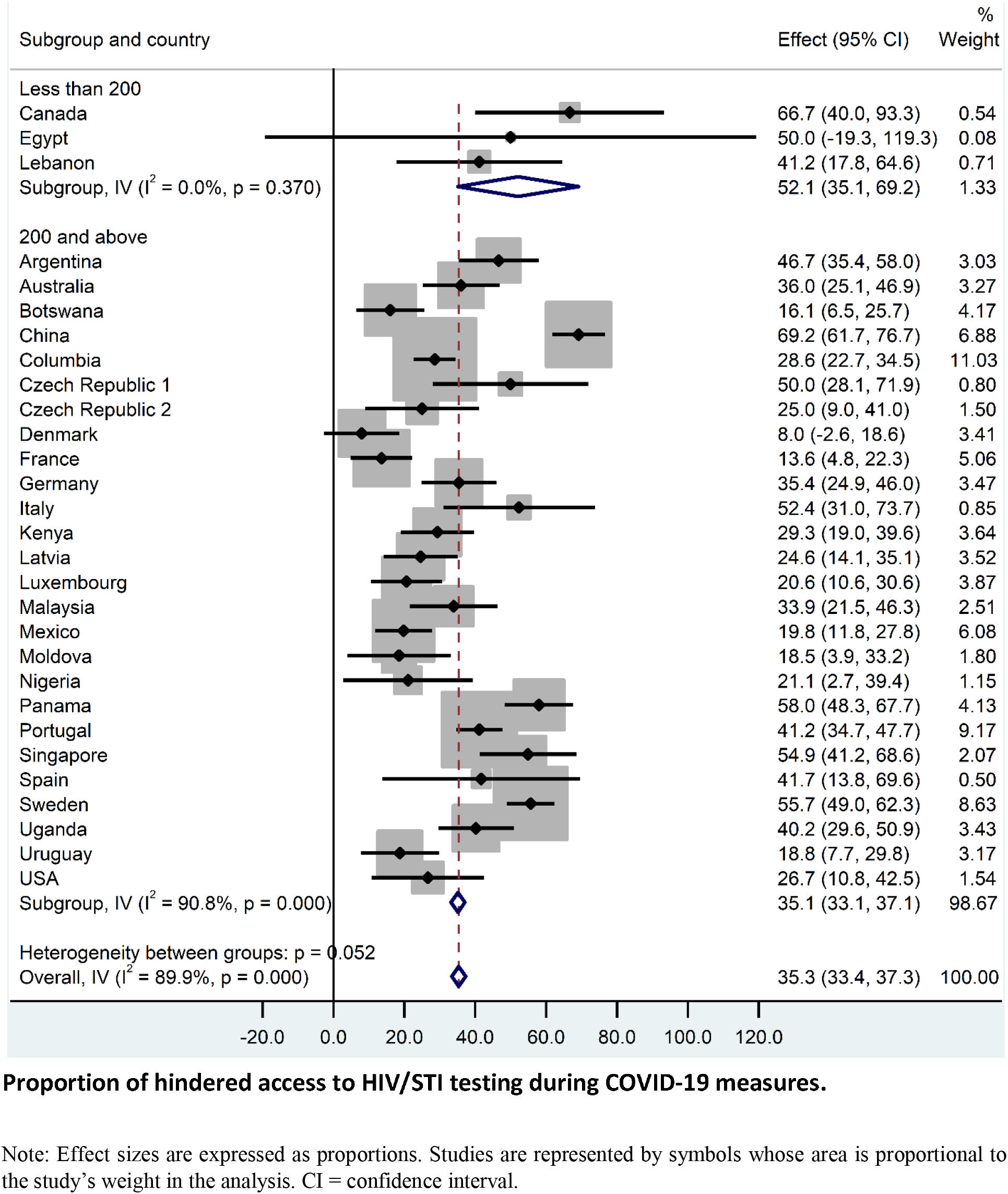
Forest plot for random-effects meta-analysis of the association between sample size and access to HIV/STI testing during COVID-19 measures

**Supplemental Figure 3.**
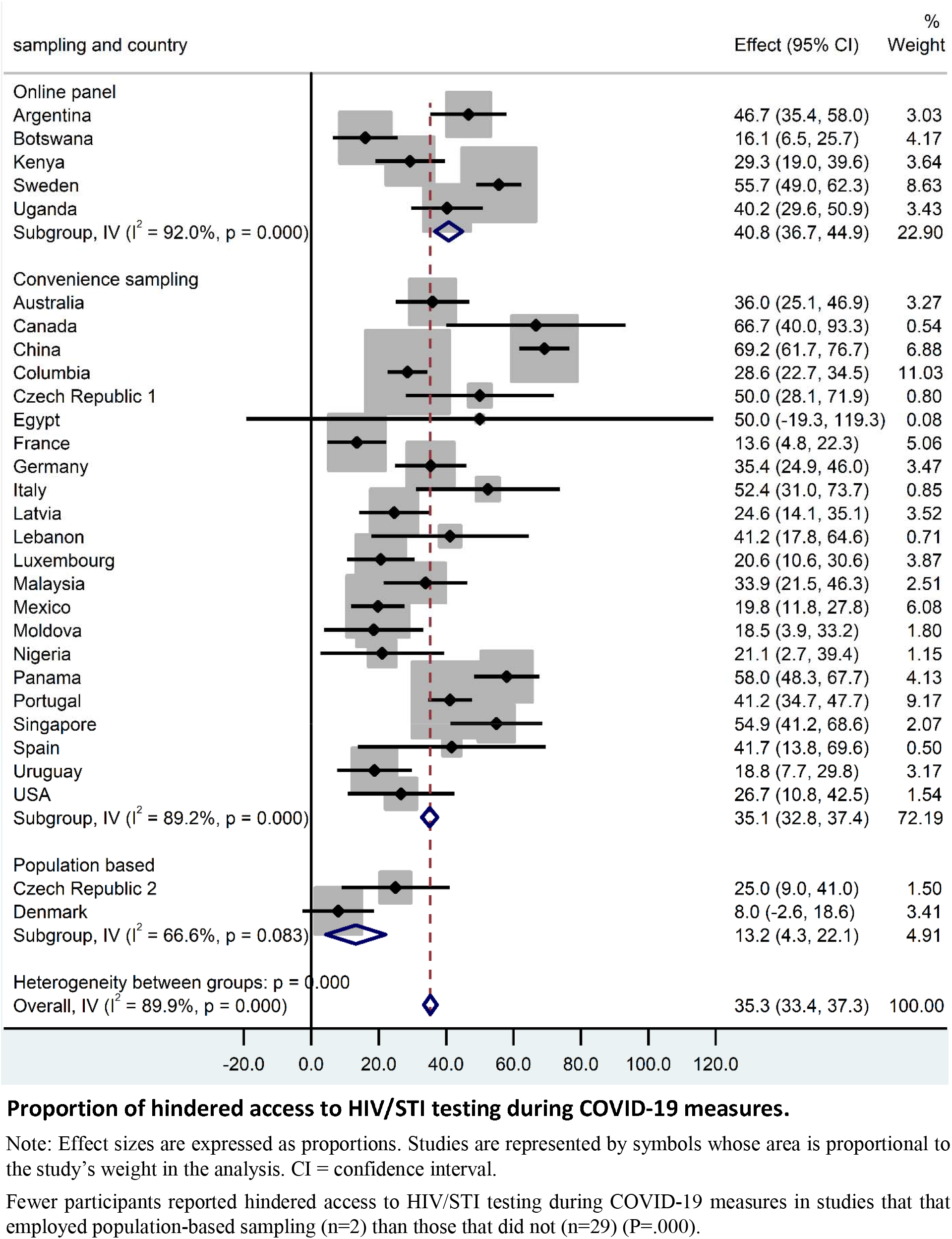
Forest plot for random-effects meta-analysis of the association between sampling method and access to HIV/STI testing during COVID-19 measures

**Supplemental Figure 4.**
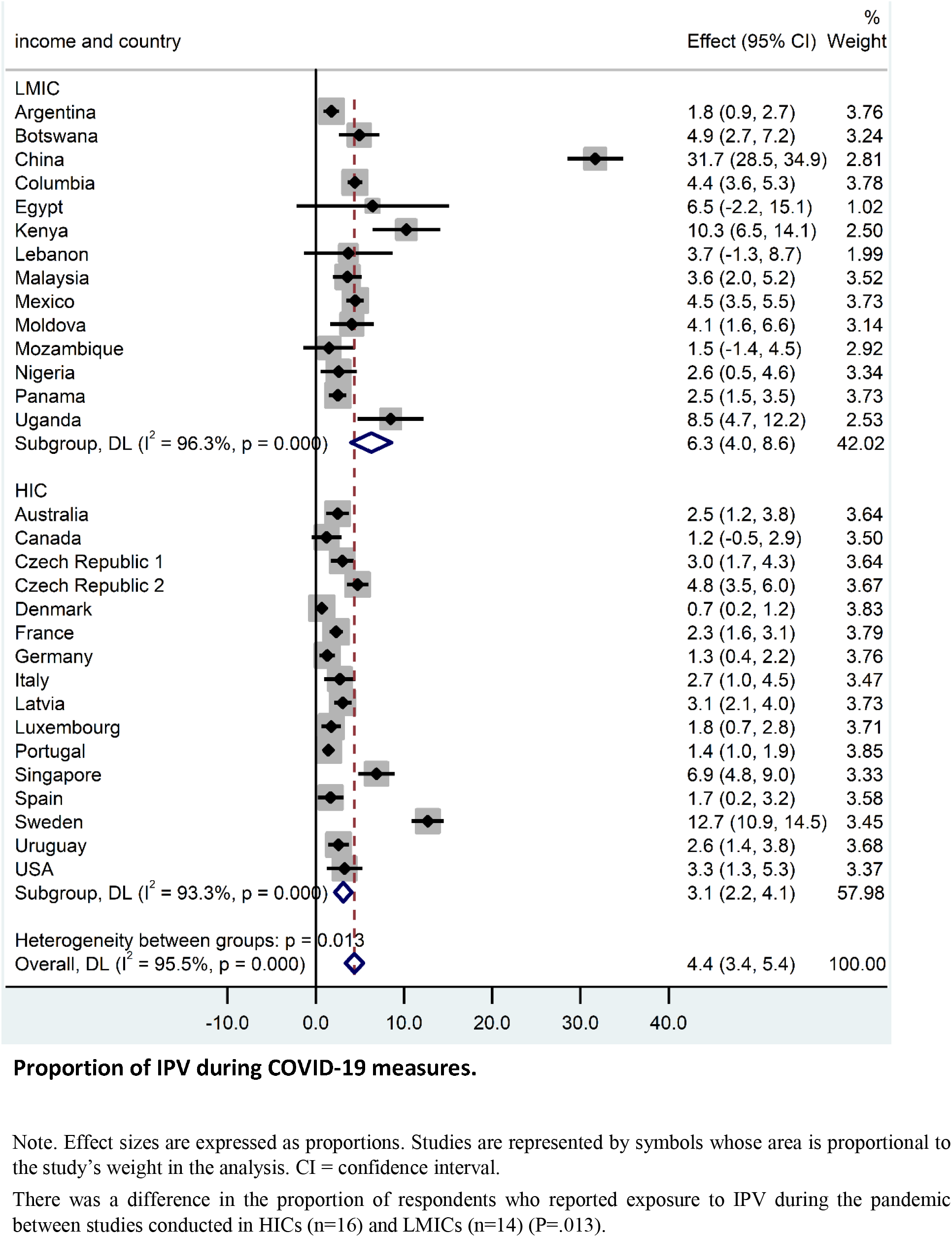
Forest plot for random-effects meta-analysis of the association between country income level and exposure to IPV during COVID-10 measures

**Supplemental Figure 5.**
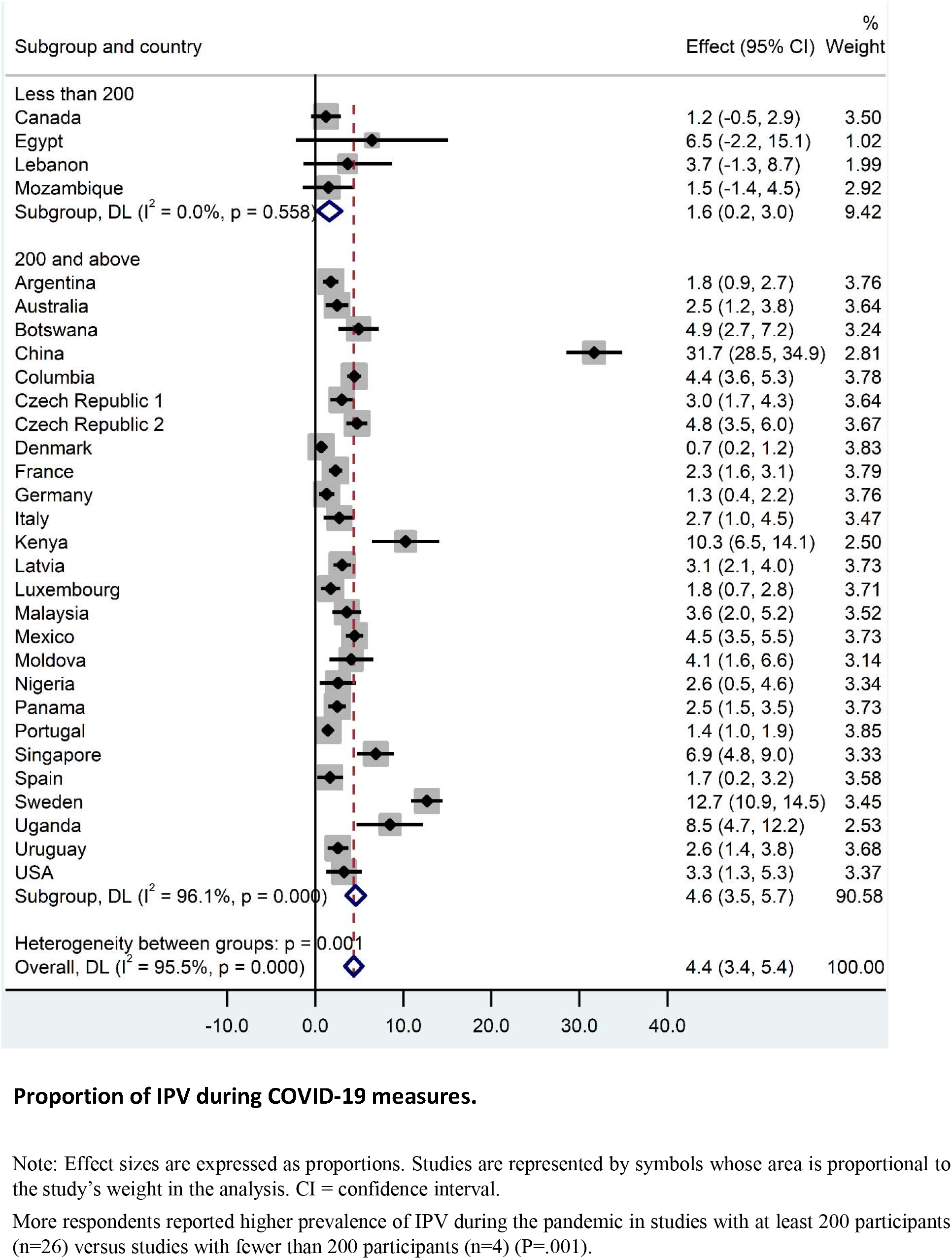
Forest plot for random-effects meta-analysis of the association between sample size and exposure to IPV during COVID-19 measures

**Supplemental Figure 6.**
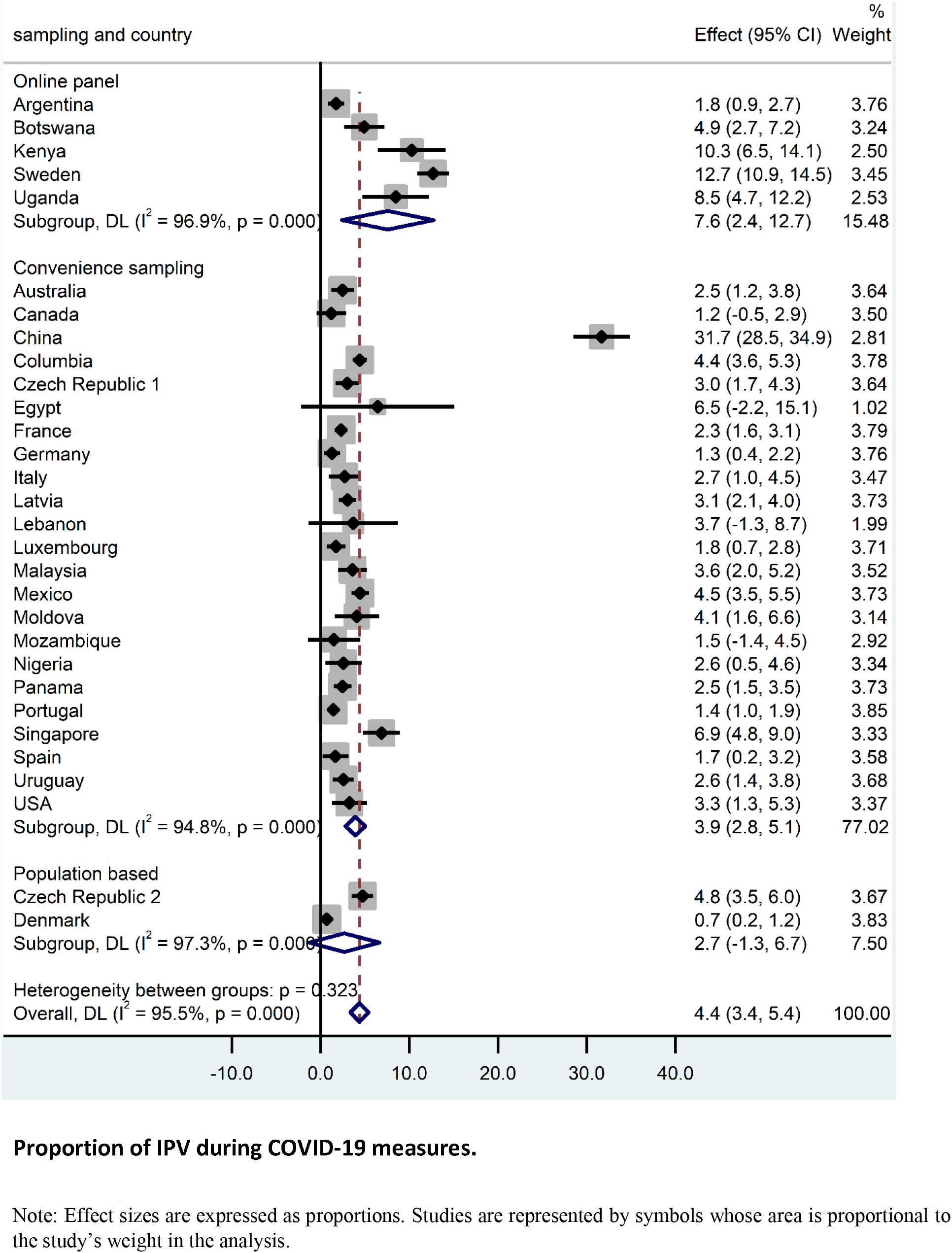
Forest plot for random-effects meta-analysis of the association between sampling method and exposure to IPV during COVID-19 measures

**Supplemental Figure 7.**
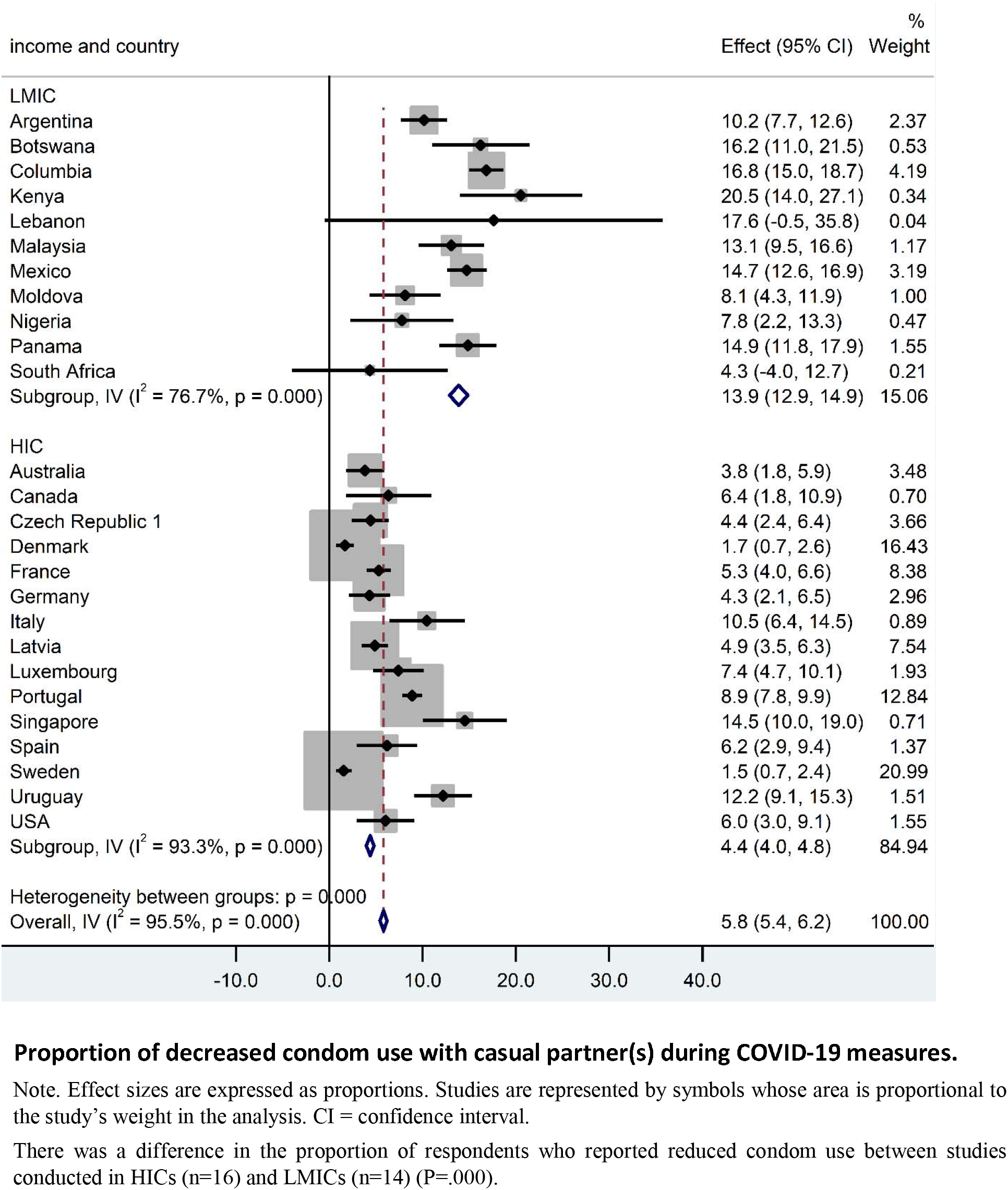
Forest plot for random-effects meta-analysis of the association between country income level and decreased condom use with casual partner(s) during COVID-10 measures

**Supplemental Figure 8:**
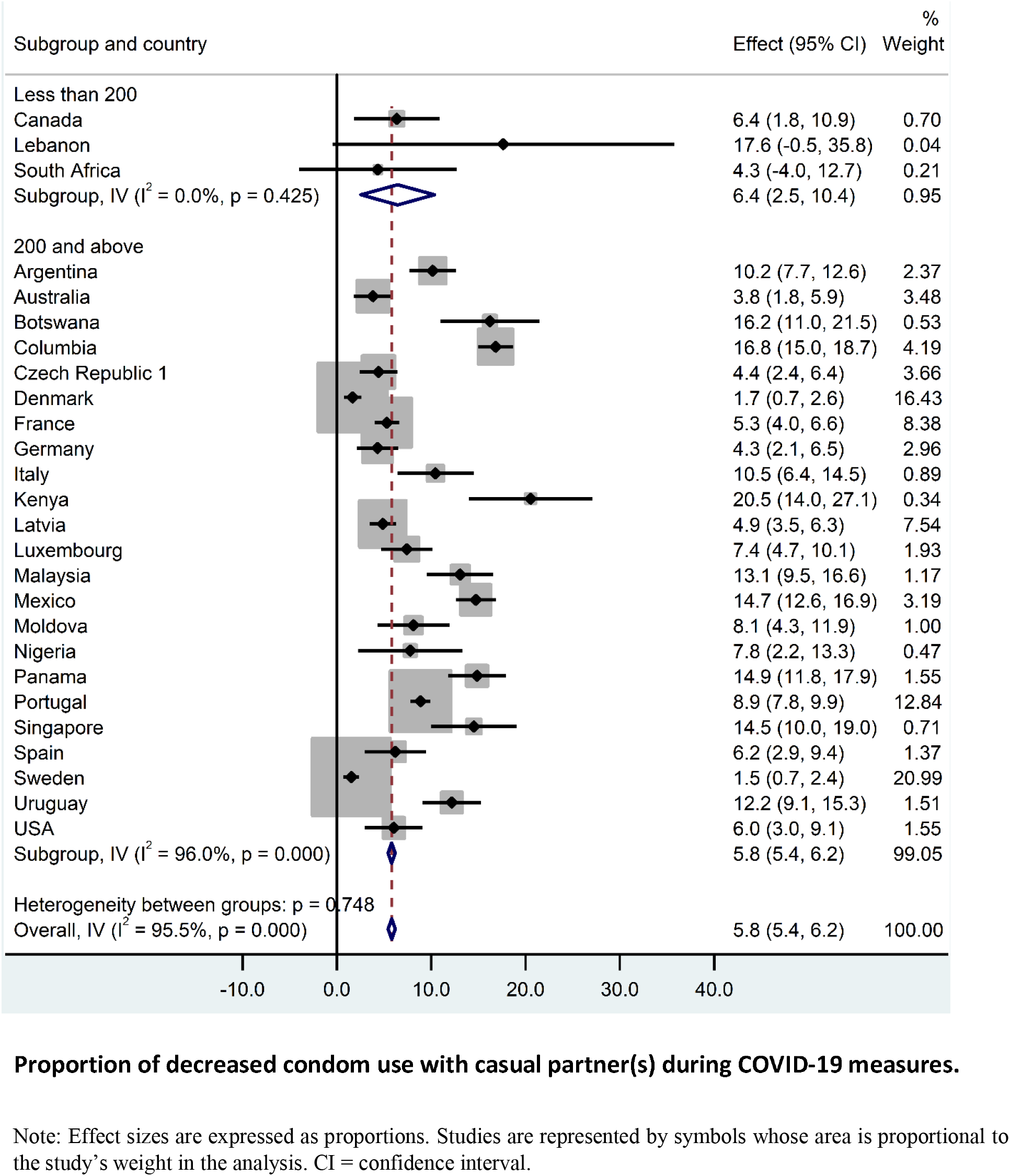
Forest plot for random-effects meta-analysis of the association between sample size and decreased condom use with casual partner(s) during COVID-10 measures

**Supplemental Figure 9:**
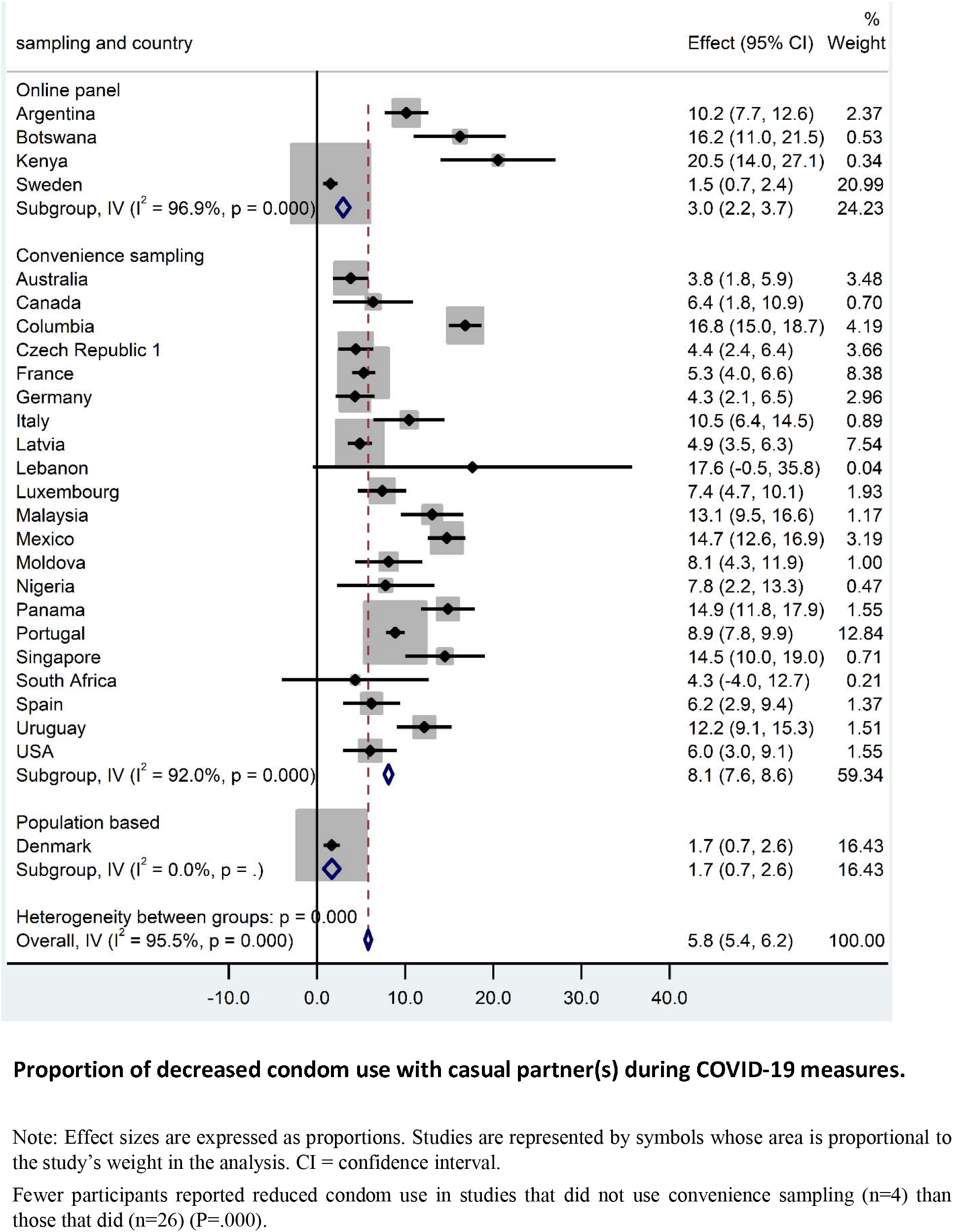
Forest plot for random-effects meta-analysis of the association between sampling method and decreased condom use with casual partner(s) during COVID-10 measures

